# Environmental hazards from pollution of antibiotics and resistance-driving chemicals in urban river networks of Malawi

**DOI:** 10.1101/2025.04.15.25325896

**Authors:** Derek Cocker, Taonga Mwapasa, Roman Grabic, Kateřina Grabicová, Andrea Vojs Staňová, Kondwani Chidziwisano, Adam Roberts, Tracy Morse, Nicholas Feasey, Andrew Singer

## Abstract

African communities have a high prevalence of antimicrobial-resistant bacterial carriage, alongside high levels of antibiotic usage and environmental pollution. Limited access to water, sanitation and hygiene infrastructure and wastewater treatment facilities enables the dissemination of resistant bacteria, antimicrobials and antibiotic resistance-driving chemicals (ARDCs) into local rivers. Few data exist quantifying the chemical drivers of antimicrobial resistance (AMR) in urban aquatic environments from African settings. In this longitudinal surveillance study, we investigated urban rivers in Blantyre, Malawi over an uninterrupted 12-month period, identifying a broad-range of chemical pollutants in urban river systems, including antibiotics, common pharmaceuticals, agricultural and industrial chemicals and heavy metals. Antimicrobial concentrations were found at levels selective for AMR and ARDCs exhibited seasonal variations, indicating that deficient sanitation infrastructure and anthropogenic factors result in high antibiotic and ARDC levels entering the river systems, which serve as an important ecological niche for the acquisition, maintenance and transmission of AMR.

## Introduction

Antibiotics are primarily used in the treatment and prevention of disease in humans and animals, alongside the promotion of growth within the animal sector (1). Antibiotic resistance (AMR) is annually associated with 4.95 million human deaths and will lead to an estimated economic loss of US$100 trillion every year by 2050 if urgent action is not taken (2,3). Global health inequities and limited absence of access to reserve antibiotics mean that the greatest burden of AMR will be felt in low and middle-income countries (LMICs) (2,4). In these settings, AMR poses an additional threat to the livestock sector and, thus, to the livelihoods of millions who raise animals for subsistence (5).

The role of the environment as a reservoir for resistant pathogens has increasingly become clear, with a growing evidence base for its relevance to human health (6). As such, it is critical to adopt a One-Health approach when considering interventions that tackle AMR on a global scale (7,8). Around 40-90% of antibiotics consumed by humans and animals are excreted in an active form, and these are dispersed into groundwater and the wider riverine network in the absence of adequate sanitation infrastructure (7,9). The presence of antibiotics, alongside other key antibiotic resistance-driving chemicals (ARDCs) (i.e., pesticides and metals) in aquatic environments, promotes horizontal gene transfer and alters microbial communities, contributing to the dissemination of antimicrobial-resistance genes (ARGs) and subsequently poses downstream risks to human, animal, and ecological health (7,10–12). In certain settings, this is compounded by pollution from inadequate treatment of healthcare-associated, industrial, domestic, and agricultural waste, boosting the xenobiotic-derived resistome in the environment (13).

Within African countries, it is frequently still the case that there is a paucity of adequate water, sanitation and hygiene (WASH) infrastructure; instead WASH behavioural practices that lead to high levels of faecal contamination of local rivers are prevalent, including the use of these open waters for domestic purposes (14,15). Increasing urbanisation within African countries poses further health hazards via the pooling of domestic sewerage and agricultural run-off from subsistence and small-scale farming (14,16). Many African countries are prone to extreme seasonal changes in rainfall and temperature. This climate exacerbates the risk of untreated sewage entering the environment through heavy rainfall, overwhelming the sewage network and promoting the survival and dissemination of human enteric pathogens. Research on the distribution and ecological hazards posed by resistance-driving chemicals and AMR bacteria and genes in urban rivers from these settings is scarce, particularly in sub-Saharan Africa (sSA) (17). Therefore, it is important to establish a baseline for the presence of antibiotic residues and co-selecting agents (e.g., pharmaceuticals, herbicides, insecticides, metals) from waterways (9,12). This knowledge could be used to conduct hazard assessments for human and environmental exposure to AMR bacteria, genes and the resistance-driving chemicals themselves. Moreover, knowledge of the chemical state of rivers is required to gauge the success of future interventions and stewardship efforts aimed at reducing the AMR burden in Africa.

Here, we investigate the presence of and fluctuations in river water contamination with metals and ARDCs, including human and animal medications, insecticides, herbicides and fungicides, at key sites within the urban riverine network of Blantyre, Malawi. Additionally, we evaluate seasonal variations in antibiotic concentrations and quantify ecological risks of AMR using predicted no-effect concentration (PNEC) thresholds agreed by the AMR Industry Alliance for antibiotic discharge to the river environment (10).

## Results

Sites in the urban river systems of Blantyre, Malawi were screened for acceptability to the populace and technical feasibility of sampling (**Table S1**) with 2 sites selected for longitudinal surveillance over an uninterrupted 12-month period between November 2020 to November 2021 (**Figure 1**). Site 1 is a river location downstream of the city centre/hospital and site 2 is a river location downstream of a dense urban community (**Figure 1b)**. Both sites were found to be heavily contaminated with ARDCs, including medications intended for human use alongside products typically used in agriculture. In total, 49 pharmaceuticals, 38 antibiotics, 8 antiretrovirals, 2 antifungals, 3 antiparasitics, 10 insecticides, 28 herbicides, 3 industrial chemicals, 8 fungicides and 25 heavy metals were recovered from rivers in urban communities (**Figures S1 & S2a-j**).

**Figure 1.**
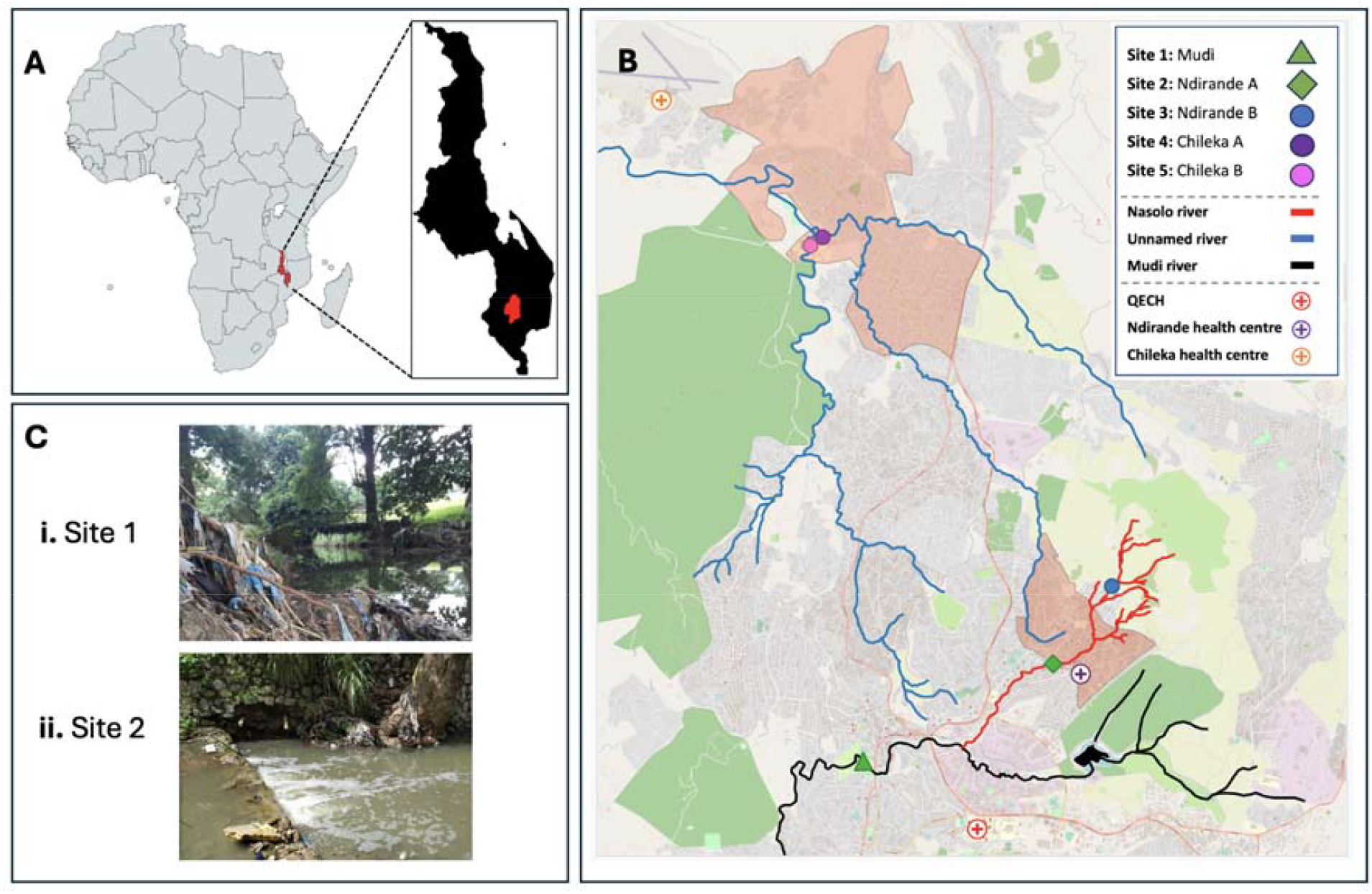
Description of the study setting, riverine networks and sampling sites. Sampling was undertaken in Blantyre city in southern Malawi (panel A). 5 river locations were screened during the pilot phase, including the Mudi river downstream of the urban centre (site 1), the Nasolo river, below (site 2) and above (site 3) the Ndirande township (shaded orange) and at two points along an unnamed river that flow through peri-urban communities on the outskirts of the city (sites 4 and 5 in Chileka, shaded orange). Sites 1 & 2 were enrolled into the longitudinal surveillance study, based on consistent year-round flow, logistics and safety profiling **(appendix Table S1 & Figures S13-5)** and photos of these river sites at initiation are seen in Panel C (Ci = Site 1, Cii = Site 2).

### Metals

Twenty-five different metals were repeatedly found in the rivers via grab samples (n=55, Site 1: n=27, Site 2: n=28) taken at weekly intervals over 6 months, with only 2 metals below the limit of quantification across all sites (Be and Sn) (**Table S2**). Metal concentrations (µg/L) varied by site and element, with median concentrations of Cu, Cr, Fe, Ni, Sb and Zn shown to be higher in the central urban river system downstream of the city centre (**Figure 2**, site 1) and metal concentrations of As, Li, Rb and Sr higher in the river systems downstream of the dense urban conurbation (**Figure 2**, site 2). Whilst none of the median (IQR) concentrations exceeded recognised World Health Organisation (WHO) or United States Environmental Protection Agency (USEPA) water quality standards (18), isolated high levels of Ni (>20µg/L), Mg (>100µg/L) and Fe (>300µg/L) were recorded in excess of these levels (**Figure 2 & Table S2**).

**Figure 2.**
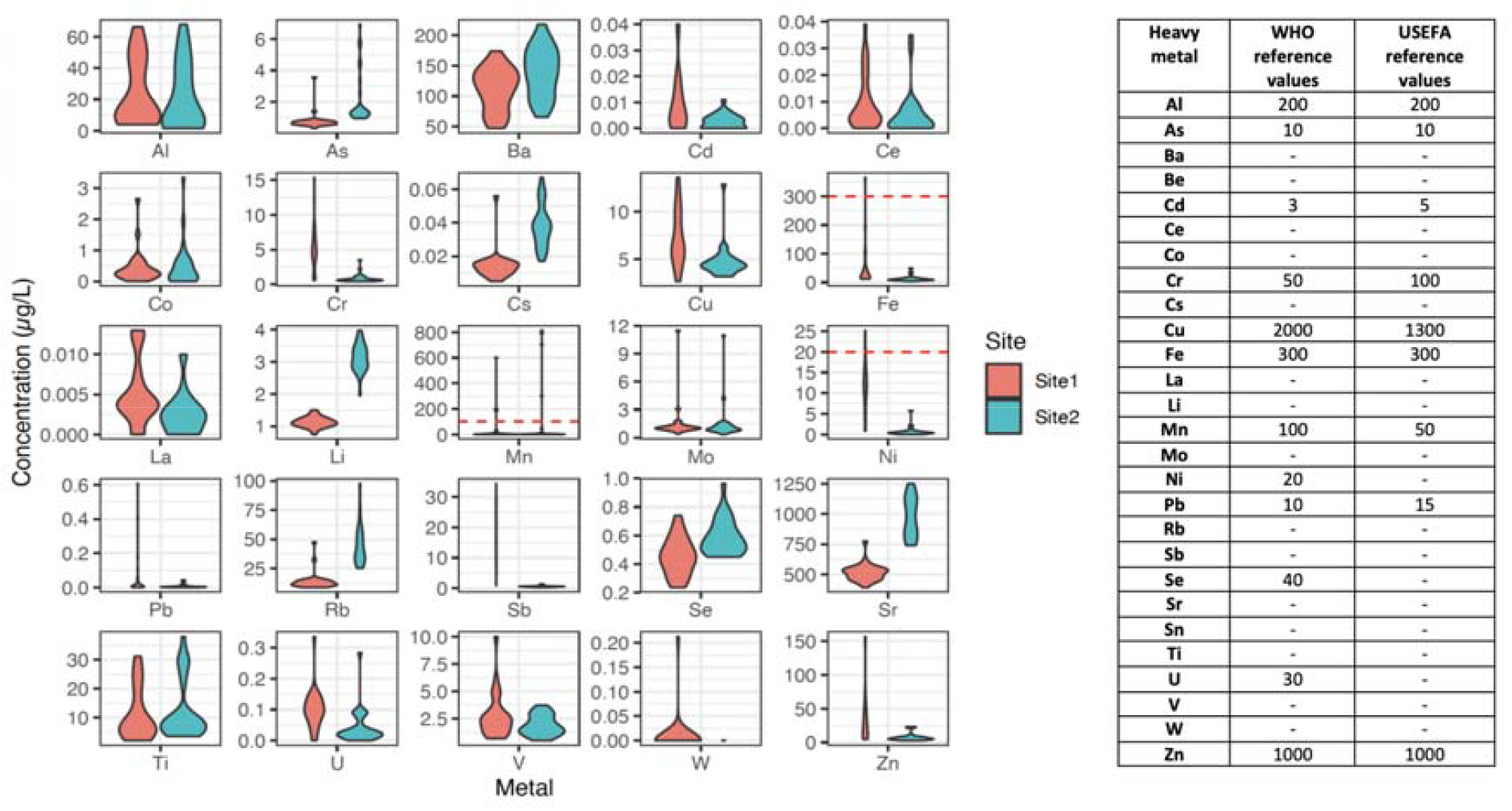
The presence and variations in heavy metals concentrations (µg/L) identified in urban waterways. (A) Violin plots of heavy metal concentrations, stratified by site, including where acceptable levels have been exceeded by WHO reference standards [denoted by a dashed red line]. (B) A list of accepted international reference standards for heavy metal concentrations (WHO / USEFA).

### Resistance-driving chemicals and medications

Spatiotemporal variations in ARDCs were found in urban rivers, with insecticides, herbicides and fungicides exhibiting fluctuating levels throughout the year (**Table 1 & Figures S3a-b**), in contrast to antibiotics and human medications, which were often seen at consistently elevated levels (**Table 2 & Figures S3a-b, S4a-b & S5a-b**). Principle Component Analysis (PCA) highlighted that chemical composition differed substantially between sites, likely reflecting differences in the geography upstream of the rivers (light industry and tertiary hospital effluent vs dense conurbation and agricultural land) (**Figure S1**).

**Table 1.** Anthropogenic pollution of urban rivers with insecticides, herbicides, fungicides and industrial chemicals. Insecticides, herbicides, fungicides and industrial chemicals recovered via continuous sampling of rivers in urban Blantyre over a 1-year period are presented with their mean (SD) concentrations (ng.POCIS^-1^.day^-1^). Differences in the concentrations of chemicals seen at sites (1-2) or between seasons (wet-dry) have been highlighted (Man-Whitney (site) Wilcoxon test (season) p<0.05 [blue = site, green = season]). The distribution of chemical concentrations (ng.POCIS^-1^.day^-1^) are included in the supplementary information (**Figures S6a-d)**. Chemical metabolites = *. Chemicals where the limit of quantification (<LOQ) was not reached as shown.

|  | Chemical compound | Site 1 mean (SD)<br>ng.POCIS<sup>-1</sup>.day<sup>-1</sup> | Site 2 mean (SD)<br>ng.POCIS<sup>-1</sup>.day<sup>-1</sup> | Site 1 ratio (pos:neg) | Site 2 ratio (pos:neg) | Site difference | Seasonal difference Site 1 | Seasonal difference Site 2 |
| --- | --- | --- | --- | --- | --- | --- | --- | --- |
| **Insecticides** | Carbofuran-3-hydroxy* | 100 (250) | 23 (13) | 42:4 | 47:3 | 0.1107 | 0.0000 | 0.0220 |
|  | Chlorpyrifos | 39 (33) | 59 (40) | 46:0 | 48:2 | 0.0033 | 0.0001 | 0.1371 |
|  | DEET | 280 (333) | 260 (180) | 46:0 | 49:1 | 0.6308 | 0.0100 | 0.2885 |
|  | Diazinon | 0.94 (0.95) | 3.1 (5.7) | 20:26 | 47:3 | 0.0002 | 0.9623 | 0.2911 |
|  | Dimethoate | 6.6 (5.7) | 11 (4) | 4:42 | 5:45 | 0.3913 | NA | NA |
|  | Imidacloprid | 23 (47) | 57 (58) | 45:1 | 48:2 | 0.0000 | 0.0119 | 0.0140 |
|  | Malathion | 2.9 (1.5) | 9.0 (6.8) | 10:36 | 30:20 | 0.0045 | 0.3608 | 0.1888 |
|  | Pirimiphos_methyl | 5.2 (5.2) | 15 (13) | 45:1 | 49:1 | 0.0000 | 0.1583 | 0.2982 |
|  | Thiamethoxam | 1.3 (0.3) | 2.8 (2.9) | 9:37 | 20:30 | 0.0054 | NA | NA |
|  | 3-chloro-4-methylaniline | 0.80 (0.56) | <LOQ | 4:42 | 0:50 | NA | NA | NA |
| **Herbicides** | Acetochlor | 7.4 (8.7) | 3.0 (2.5) | 21:25 | 33:17 | 0.1431 | 0.3915 | 0.0008 |
|  | Acetochlor_ESA* | 3.9 (3.7) | 2.2 (1.7) | 37:9 | 38:12 | 0.0424 | 0.0078 | 0.0241 |
|  | Ametryn | 0.33 (0.19) | <LOQ | 15:31 | 0:50 | NA | 0.0250 | NA |
|  | Atrazine | 0.95 (0.75) | 1.1 (1.5) | 25:21 | 34:16 | 0.8902 | 0.0046 | 0.0017 |
|  | Atrazine_desisopropyl* | <LOQ | 6.0 (3.0) | 0:46 | 3:47 | NA | NA | NA |
|  | Atrazine_desethyl-desisopropyl* | 18 (12) | <LOQ | 10:36 | 0:50 | NA | 0.2386 | NA |
|  | Atrazine_desethyl* | 46 (50) | 18 (11) | 32:14 | 26:24 | 0.1616 | 0.0050 | 0.0483 |
|  | Atrazine_2-hydroxy* | 0.78 (0.69) | 0.43 (0.42) | 42:4 | 25:25 | 0.0284 | 0.0115 | 1.0000 |
|  | Bensulfuron_methyl | 1.3 (0.4) | 0.70 (0.00) | 2:44 | 1:49 | NA | NA | NA |
|  | Clomazone | 2.2 (1.5) | <LOQ | 2:44 | 0:50 | NA | NA | NA |
|  | Diuron | 9.0 (12.1) | 6.7 (7.0) | 42:4 | 47:3 | 0.6720 | 0.1658 | 0.0366 |
|  | Diuron_desmethyl* | 1.6 (1.3) | 1.5 (0.55) | 11:35 | 5:45 | 0.1721 | NA | NA |
|  | Fluazifop-p | 0.15 (0.03) | 1.8 (1.0) | 4:42 | 3:47 | 0.0497 | NA | NA |
|  | Hexazinone | 0.20 (0.05) | 0.35 (0.15) | 7:39 | 5:45 | 0.0735 | NA | NA |
|  | Ioxynil | <LOQ | 0.39 (0.00) | 0:46 | 1:49 | NA | NA | NA |
|  | MCPA | 4.3 (4.9) | 3.6 (2.7) | 4:42 | 2:48 | 0.817 | NA | NA |
|  | Metazachlor | 5.7 (0) | 0.14 (0.00) | 1:45 | 1:49 | NA | NA | NA |
|  | Metolachlor | 4.3 (4.9) | 1.5 (0.7) | 46:0 | 48:2 | 0.0013 | 0.5164 | 0.0185 |
|  | Metolachlor_ESA* | 8.2 (8.3) | 2.3 (1.3) | 43:3 | 29:21 | 0.0000 | 0.4144 | 0.8318 |
|  | Metribuzin_desamino* | 0.94 (0.00) | <LOQ | 1:45 | 0:50 | NA | NA | NA |
|  | Picloram | 5.7 (0.0) | 6.8 (2.5) | 1:45 | 10:40 | NA | NA | NA |
|  | Prometryn | 4.3 (3.7) | <LOQ | 17:29 | 0:50 | NA | 0.4615 | NA |
|  | Propazine_2-hydroxy* | <LOQ | 0.95 (0.00) | 0:46 | 1:49 | NA | NA | NA |
|  | Simazine_hydroxy* | 0.53<br>(0.40) | 1.2 (1.1) | 29:17 | 30:20 | 0.0129 | 0.8384 | 0.0513 |
|  | Terbutylazine | 0.25<br>(0.12) | 0.42<br>(0.19) | 21:25 | 29:21 | 0.0001 | 0.2806 | 0.2290 |
|  | Terbutylazine_hydroxy* | 0.71<br>(0.44) | 0.52<br>(0.23) | 36:10 | 33:17 | 0.1615 | 0.0960 | 0.0497 |
|  | Terbutryn | 0.96<br>(0.70) | 1.4 (0.9) | 43:3 | 47:3 | 0.0059 | 0.4123 | 1.0000 |
|  | 1-(3,4-Dichlorophenyl)_urea* | 0.99<br>(0.41) | 3.2 (3.4) | 9:37 | 14:36 | 0.0039 | NA | NA |
| Industrial chemicals | 1H-benzotriazol | 630<br>(1520) | 100 (95) | 46:0 | 48:2 | 0.0007 | 0.0731 | 0.4767 |
|  | 1H-benzotriazol_(5/4)-methyl* | 92 (116) | 20 (26) | 44:2 | 44:6 | 0.0001 | 0.0000 | 0.0000 |
|  | 1H-benzotriazol_1-methyl* | 1500<br>(1500) | 560<br>(690) | 26:20 | 24:26 | 0.0296 | 0.3606 | 0.1299 |
| Fungicides | Azoxystrobin | 0.69<br>(0.68) | 0.53<br>(0.19) | 25:21 | 29:21 | 0.8215 | 0.0457 | 0.5354 |
|  | Carbendazim | 6.4 (5.5) | 17 (12) | 45:1 | 49:1 | 0 | 0.0014 | 0 |
|  | Dimethomorph | 1.3 (0.4) | <LOQ | 5:41 | 0:50 | NA | NA | NA |
|  | Epoxiconazole | 0.18<br>(0.06) | 0.46<br>(0.31) | 6:40 | 10:40 | 0.1035 | NA | NA |
|  | Flusilazole | 0.90<br>(0.00) | 0.25<br>(0.11) | 1:45 | 18:32 | NA | NA | NA |
|  | Metalaxyl | 2.0 (1.1) | 2.3 (1.2) | 3:43 | 9:41 | 0.6433 | NA | NA |
|  | Propiconazole | 0.35<br>(0.34) | 1.4 (1.1) | 42:4 | 48:2 | 0.0000 | 0.0003 | 0.0000 |
|  | Tebuconazole | 1.2 (1.1) | 3.5 (2.0) | 24:22 | 35:15 | 0.0000 | 0.5099 | 0.0001 |

**Table 2.**
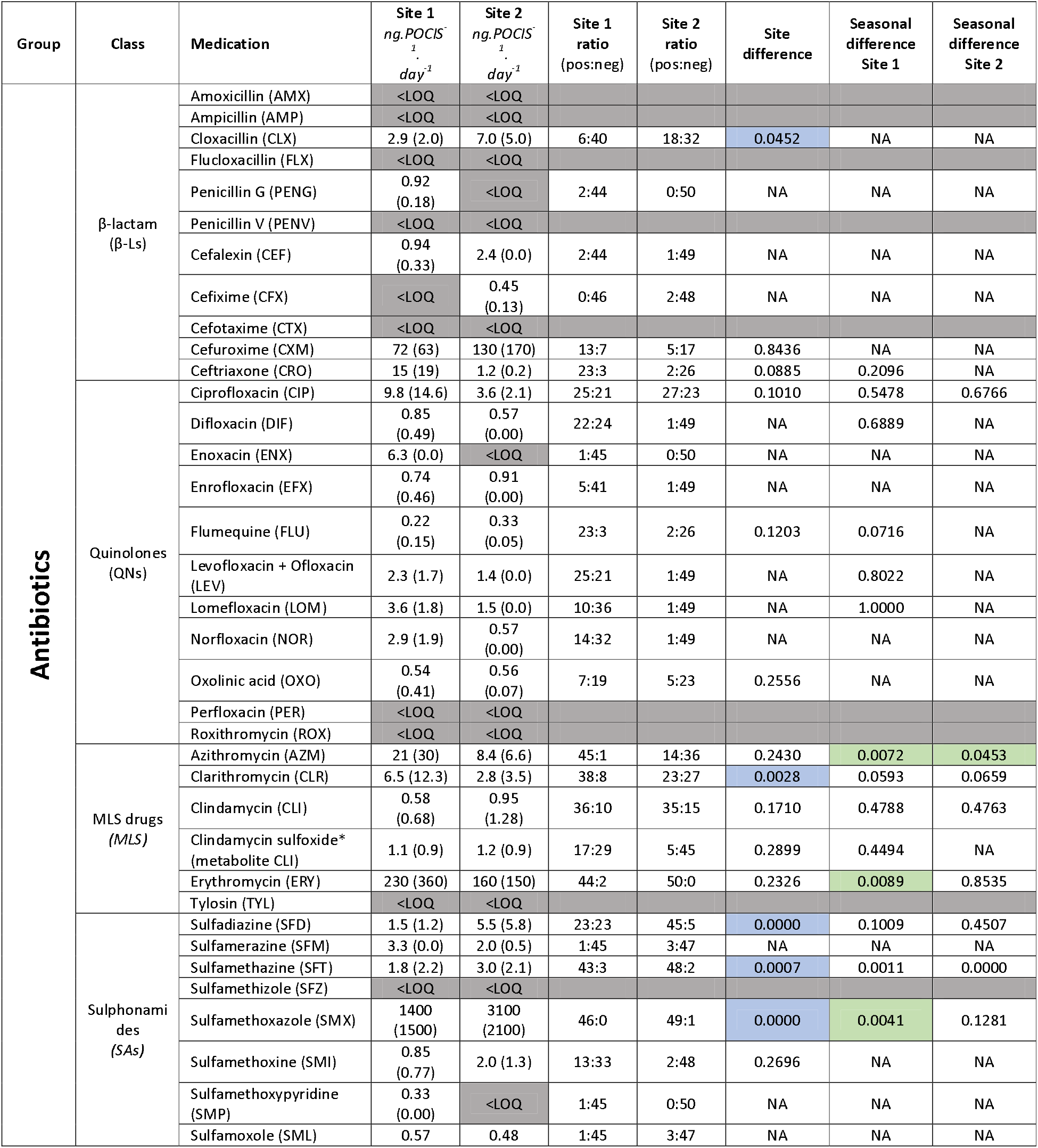
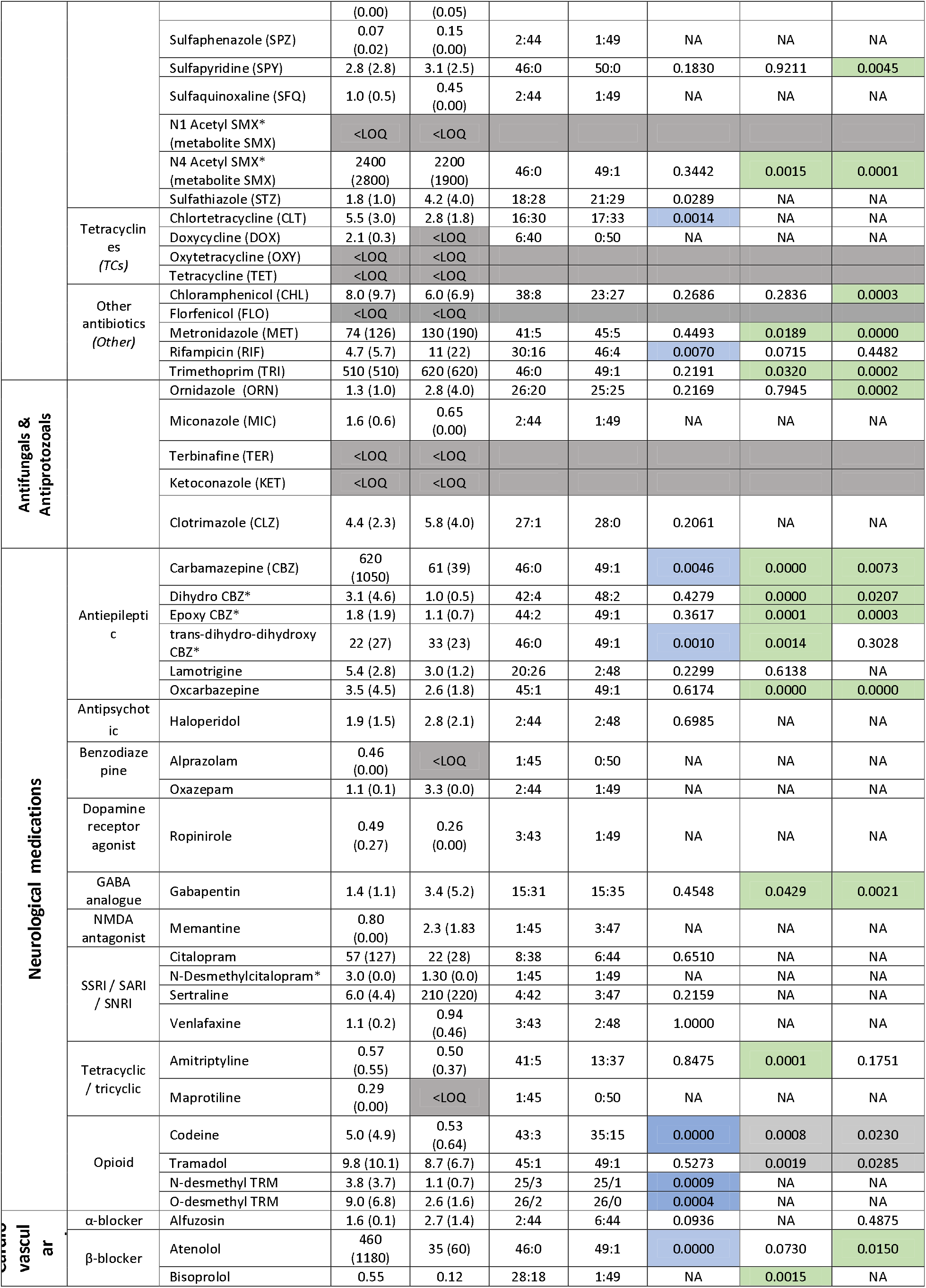
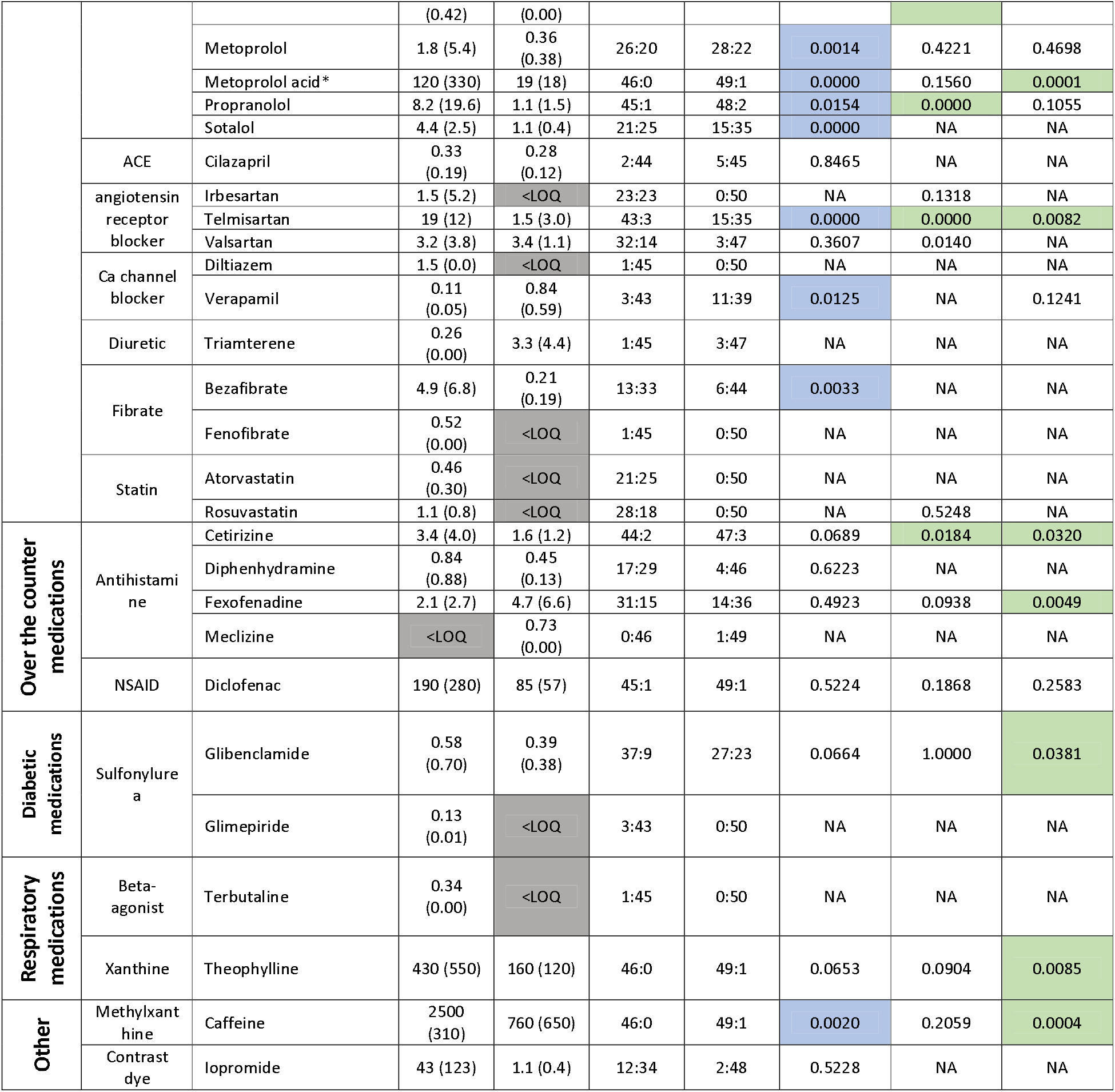
Anthropogenic pollution of urban rivers with antibiotics, antifungals, antiprotozoals and other human-use medications. Medications recovered via continuous sampling of rivers in urban Blantyre over a 1-year period are presented with their mean (SD) concentrations (ng.POCIS-1.day-1). Differences in the concentrations of medications seen at sites (1-2) or between seasons (wet-dry) have been highlighted (Man-Whitney (site) Wilcoxon test (season) p<0.05 [blue = site, green = season]). The distribution of medication concentrations (ng.POCIS^-1^.day^-1^) are included in the supplementary information (**Figures S6e-g)**. Chemical metabolites = *. Chemicals where the limit of quantification (<LOQ) was not reached as shown.

Overall, there were high levels of river contamination with chemicals used in agricultural and industrial practices (**Table 1**). 80.4% (n=37/46) of analytes were recovered from both rivers, with a detection frequency of 90.0% (n=9/10) for insecticides, 75.0% (n=21/28) for herbicides and 87.5% (n=7/8) for fungicides at both sites, and the minority found at a single location only (**Figures S2a-f**). The mean (SD) analyte concentration normalized to sampling time (ng.POCIS^-1^.day^-1^) varied by location (**Table 1 & Figures S6a-d**), with industrial chemicals and herbicides found at higher levels in the city centre (site 1) and neonicotinoid and organophosphate insecticides found at higher levels below the urban conurbation (site 2). Of note, DEET, chlorpyrifos, carbofuran and benzotriazole were found at particularly high concentrations (**Table 1**). Furthermore, a selection of ARDCs exhibited mean (SD) differences in concentrations depending on the season (wet vs. dry) (**Table 1**), with industrial chemicals found at proportionally higher concentrations at the end of the rainy season (**Figure S3a-b**), illustrating that seasonal changes in rainfall and local farming and agricultural practices lead to variations in the concentration of chemicals found in local rivers.

Medications used in human health were continuously recovered from urban rivers throughout the year (**Figures S4a-b & S7a-b)**, with 80.4% (n=41/51) found at both sites and the rest recovered primarily from the site downstream of the tertiary hospital (Site 1, n=9). Only sertraline and propranolol were found at levels that exceeded recognised PNEC or critical environmental concentration (CEC) targets (**Figures S8a-b**). Mean (SD) concentrations (ng.POCIS^-1^.day^-1^) differed by site (**Figure S6e-f**), with the highest levels of human pharmaceuticals frequently seen at Site 1 downstream of the local hospital (**Table 2**). Seasonal fluctuations existed (**Table 2 & Figures S7a-b**), a notable example being antiepileptics, which were found at higher river levels during the dry season (**Table 2 and Figures S7a-b**).

### Presence of anti-infective agents and associated AMR risk

Antibiotics were found in all river samples that underwent selected analysis (100%, n=96/96), including the presence of 12 sulphonamides (sulfadiazine, sulfamerazine, sulfamethazine, sulfamethoxazole, N4-acetylsulfamethoxazole, sulfamethoxine, sulfamethoxypyridine, sulfamoxole, sulfaphenazole, sulfapyridine, sulfaquinoxaline, sulfathiazole), 5 macrolides /lincosamide (azithromycin, clarithromycin, clindamycin, clindamycin sulfoxide, erythromycin), 6 β-lactams, including 4 cephalosporins (cloxacillin, penicillin-G, cefalexin, cefixime, cefuroxime, ceftriaxone), 9 fluoroquinolones (ciprofloxacin, difloxacin, enoxacin, enrofloxacin, flumequine, levofloxacin, lomefloxacin, norfloxacin, oxolinic acid) and members of 5 other antibiotic classes (chloramphenicol, metronidazole, rifampicin, trimethoprim, doxycycline), alongside 2 antifungal (clotrimazole, miconazole) and 1 antiparasitic (ornidazole) (**Table 2**). Nontargeted analysis further identified the presence of 8 antiretrovirals (abacavir, lamivudine, lopinavir, efavirenz, zidovudine, atazanavir, ritonavir and nevirapine), 2 antiparasitics used in malaria (sulfadoxine and pyrimethamine) and the tuberculosis antibiotic isoniazid (**Figure S1**). 86.8% (n=33/38) of antibiotics were recovered from both river locations, and 5 antibiotics (cefixime, doxycycline, enoxacin, penicillin-G, sulfamethoxypyridine) were identified at a single site (**Figure S2i-ii**).

Antibiotics contributed 56.8% (Site 1: 41.96%, Site 2: 75.47%) of the total cumulative chemicals (ng.POCIS-1.day-1) recovered from rivers (**Figure S9 & S6g**). The total concentrations of antibiotics recovered ranged from 0.22-22,000 ng.POCIS^-1^.day^-1^ (**Figures S10a-b**), and sulfamethoxazole (its metabolite N4-acetyl), trimethoprim, erythromycin and metronidazole were the dominant antibiotics found in river water, having both the highest detection frequency and mean (SD) concentrations (ng.POCIS^-1^.day^-1^) (**Table 2 & Figure S5a-b**). Additionally, we consistently identified macrolides and cephalosporins in urban rivers (**Figure S5a-b**).

Variations in mean (SD) antibiotic concentrations depended on the antibiotic class and river site (**Table 2 & Figure S6g**). In the upstream dense urban community, we typically found higher levels of sulphonamides and tuberculosis therapies. In comparison, higher levels of macrolide and fluoroquinolones were found in the city centre downstream of the local hospital (**Table 2, Figures S6g & 10a-b**). Unsurprisingly, sulfamethoxazole, its metabolite N4-acetyl, and trimethoprim were closely associated (**Figure S11**), reflecting their presence in the antibiotic therapy co-trimoxazole and its use in the HIV programme as co-trimoxazole preventative therapy (CPT). Similarly, co-trimoxazole was associated with rifampicin, pointing toward the link between HIV and tuberculosis therapy.

There were fluctuations in the concentrations of antibiotics seen on a month-month basis at both sites, reflecting the seasonality of infectious disease and rainfall (**Table 2 & Figures S5a-b, 10a-b**), which in turn impacts upon the selection pressures within the riverine environment. However, fewer seasonal variations were seen in antibiotic presence or concentration than in other human pharmaceuticals or agricultural chemicals.

### Ecological antimicrobial risk quantification

To determine whether antibiotic residues in urban rivers impacted on antimicrobial selection in the aquatic environment, monthly average concentrations were compared to published PNECs set out in the guidance from the AMR industry alliance; previously used in this manner in the UK (**Figure 3**) (8,10,17). Using this approach, the majority of individual antibiotic concentrations in urban rivers were below the PNEC threshold (80.95%, n=17/21). However, sulfamethoxazole, trimethoprim, metronidazole and azithromycin were frequently recovered at levels above the upper limit of PNEC values. Here, trimethoprim and metronidazole were found at ∼2 times the limit of PNECs, azithromycin was found at >3 times the PNEC and sulfamethoxazole was recovered all year round, and at levels that sometimes exceeded >10 times the PNEC threshold. Composite levels of macrolides showed additional levels of risk (**Figures S12a-b**).

**Figure 3.**
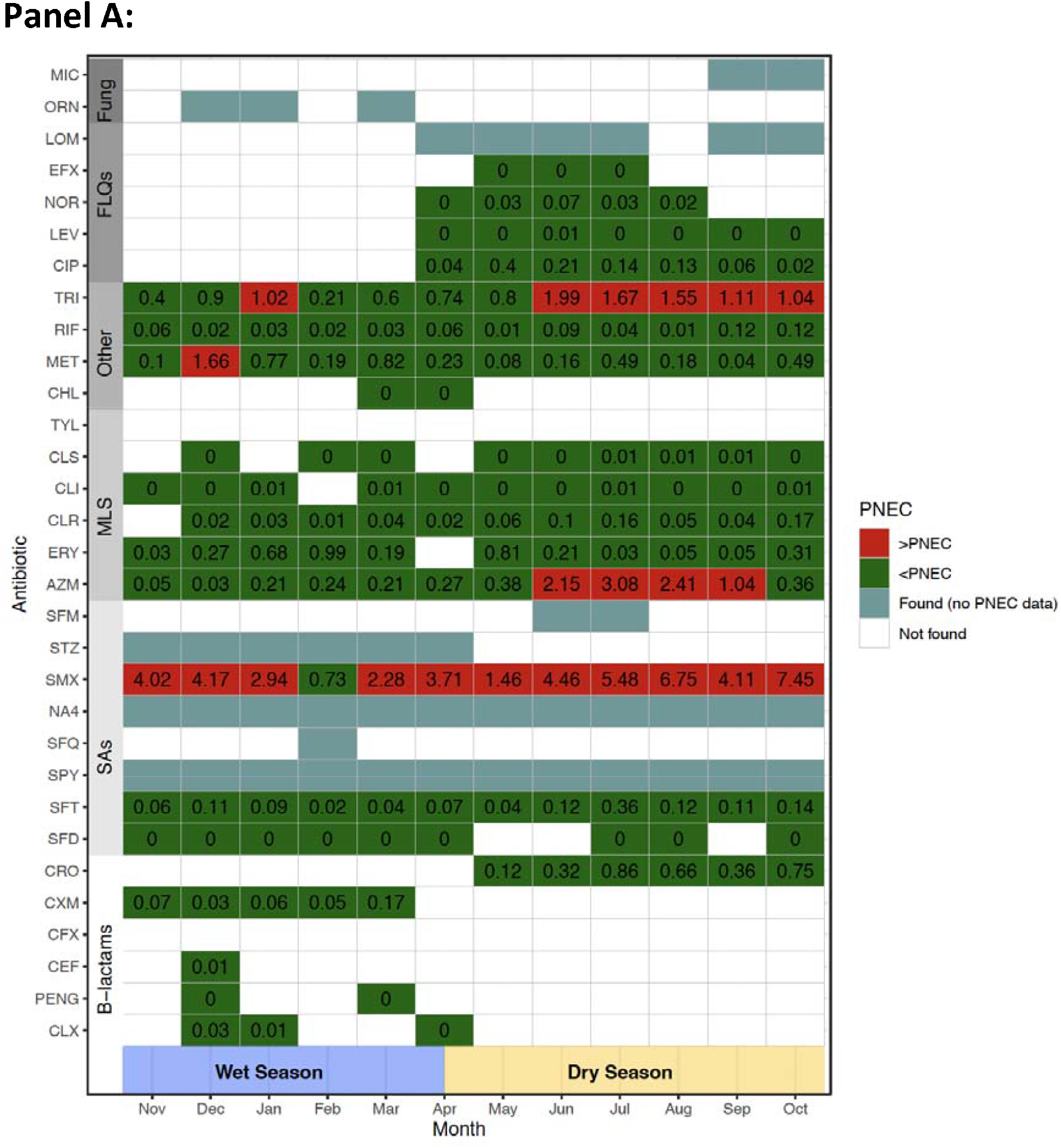
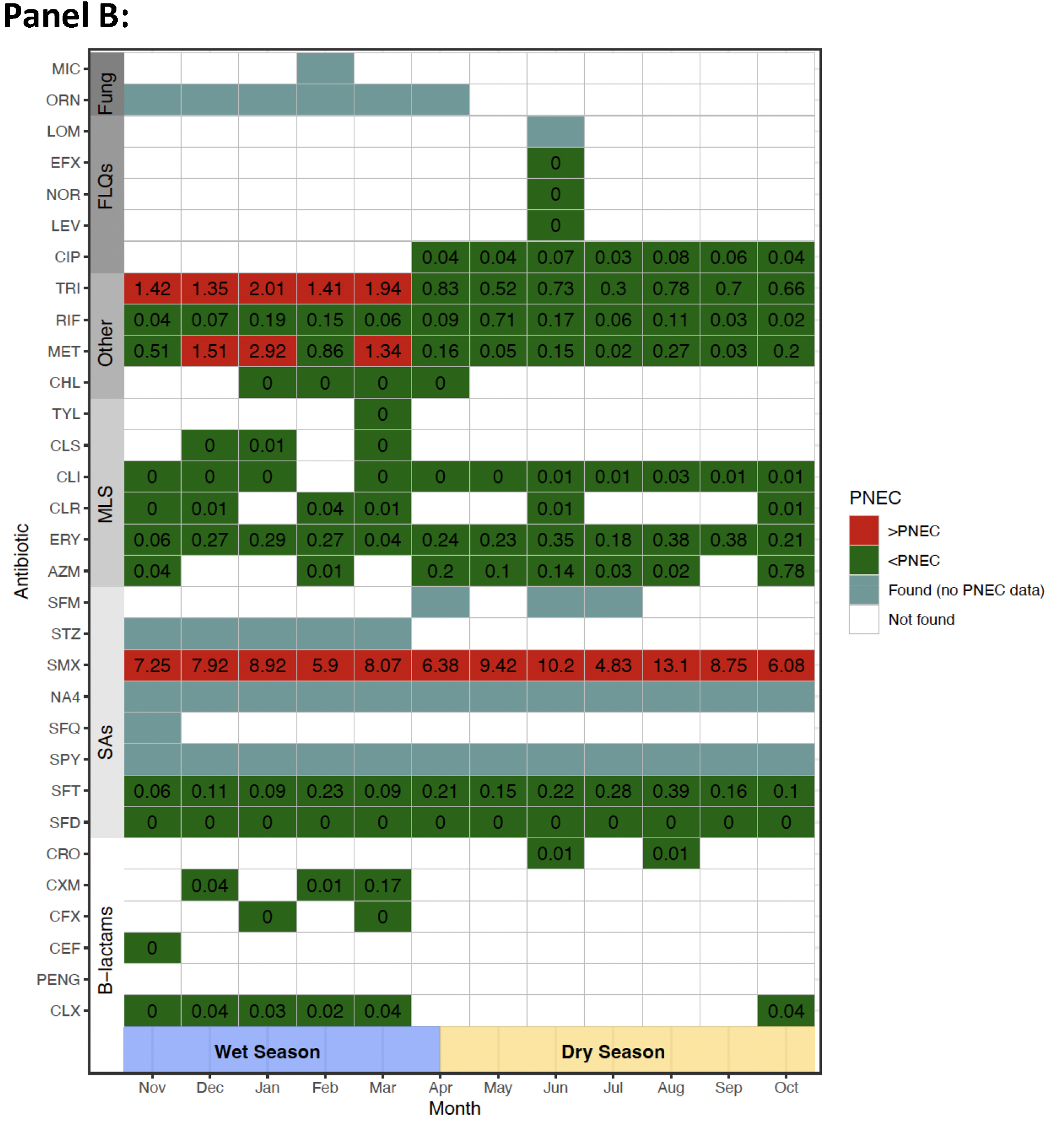
Temporal relationships in the recovery of antibiotics in river water, highlighting the continued presence of unsafe PNEC levels. Monthly trends in the presence and absence (white) of antibiotics are plotted over a 1-year period, spanning across the wet (blue) and dry (yellow) season at site 1 (A) and site 2 (B). Antibiotics are grouped by class, and stratified into safe (green, <PNEC) and unsafe (red, >PNEC) levels based on the concentrations identified. Values inside the cells describe the ratio of analyte:PNEC illustrating the levels of risk. A value of 0 denotes where an antibiotic was identified above the LOQ but below 0.01% of the agreed PNEC target. Cases where antibiotics are recovered, but there are no agreed PNEC definitions have also been highlighted (turquoise).

## Discussion

ARDC pollution into freshwater systems creates the conditions for the maintenance of intrinsic antimicrobial resistance, the selection for new resistance mutations and/or the acquisition of mobile genetic elements conferring AMR (19). The presence of antibiotics, crop and industrial chemicals, and metals in the environment increases the selection rate for antibiotic resistance, thereby allowing the environment to form a key niche for the maintenance and evolution of AMR (7,16). Here, we illustrate that ARDCs are consistently recovered from urban waterways in a large African city, including antibiotics at levels above PNECs and heavy metals above WHO reference limits, posing onward risks to human, animal, and ecological health.

Metals have previously been identified in urban sSA river systems via point prevalence studies (20). However, the longitudinal data in this study highlight that concentrations occasionally exceed putative selection thresholds, illustrating ecological risks from contamination with heavy metals fluctuates and continuous surveillance and intervention is required. Additionally, agrochemicals, including insecticides, herbicides, and fungicides frequently used by households and subsistence farmers in Africa (21) were detected and regularly recovered throughout the year. Whilst internationally agreed AMR thresholds in surface waters do not exist for these chemicals, their role is widely reported to influence selection pressures on bacteria in the aquatic environment via similar mechanisms to antibiotics, making the putative thresholds for selection (PNECs) conservative estimates of the actual minimum selective concentration (12,17,22).

Previous research in African surface waters has identified sulphonamides, such as sulfamethoxazole, as the most commonly recovered antibiotics (17,23). In this study, sulfamethoxazole was the antibiotic found at the highest concentrations, followed by trimethoprim, erythromycin/azithromycin, metronidazole and rifampicin. Despite a growing trend in intensive farming practices and the use of antibiotics for growth promotion in Malawi (24), this spectrum of antibiotics reflects those typically used locally in human health to treat a broad range of bacterial diseases (25). Human antibiotic usage in sSA is complex and influenced by the massive burden of infectious disease, vulnerabilities of access and cost and other intrinsic health system constraints (25). These conditions lead to a narrow spectrum of oral antibiotics typically being used within Malawi (15,24), reflected in antibiotic prevalence in urban surface waters. While campaigns to optimise community antibiotic prescribing are ongoing, antibiotic prescription is a rationale response in settings where the burden of infectious disease is large and diagnostics are few. Therefore, a priority focus should be on waste management improvements and environmental control of antibiotic dispersal, instead of reducing potentially life-saving antimicrobial therapy (26).

AMR selection risks posed by antibiotics and medications can be assessed using PNECs, initially proposed for use as discharge limits from manufacturing facilities or via critical environmental concentrations (10,17). When we compared these targets to antibiotic concentrations found in urban rivers, sulfamethoxazole, trimethoprim, metronidazole and azithromycin were consistently recovered above recommended PNEC limits over extended periods of time. Given the absence of antibiotic manufacturing plants or functioning WWTP upstream of the river sites, these results suggest that ineffectual waste management of human effluent leads to the widespread dissemination of antibiotics in the urban riverine environment and that antibiotics, ARDCs, and heavy metals may serve as an important driver for AMR bacteria and ARGs in urban sSA rivers (15). Tighter regulation of ARDCs and protecting urban waterways from chemical contamination may positively impact human health and the ecosystem.

Widespread co-contamination of the urban environment with ARB is commonplace. Within Blantyre, extensive soil, surface water and environmental contamination with ESBL Enterobacteriaceae (ESBL-E) is found in high-density environments, linked to several key environmental exposure risks (27). This is further exacerbated by a lack of awareness of the risks posed from environmental pathways including human-environmental interactions, especially via contaminated river water (28). A large, contemporaneous One Health study undertaken in the dense urban conurbation directly upstream of our river site(s) found extensive ESBL-E contamination within the rivers, drains and local environments, alongside high levels of ESBL-E gut colonisation of animals and humans (15). Genomic analysis of ESBL **E. coli** isolates from human/animal stool and environmental sources identified these cluster independently of ecological source (29), highlighting the importance of the shared environment in driving AMR. Taken with the results presented in this study that show high levels of antibiotic and ARDCs in the local river systems, this reaffirms that in Malawi, AMR solutions require a One Health approach (26).

Previous research conducted in Blantyre has found seasonal relationships between the wet season and human AMR colonisation (15) and environmental AMR contamination (27). The presence of ARDCs in sub-tropical rivers is also influenced by seasonal trends in rainfall, as illustrated by our findings showing fluctuations in the recovery and concentration of antibiotics and ARDCs over time, including wet vs dry seasons. High levels of rainfall leads to widespread flooding, increased runoff from agricultural sites and overflowing of pit latrines into local rivers and groundwater, increasing exposure to pathogens, ARDCs and AMR risks. The paucity of adequate sanitation infrastructure in urban settings intensifies the effects of climactic events, impacting fluctuations of antibiotics and ARDCs in effluent and the local river systems.

Many factors can impact the fate of antibiotics and ARDCs in the aquatic environment, including photodegradation, biodegradation and river flow rates. If these factors fluctuate throughout the year it could contribute to shifts in the recovery of some analytes, impacting the interpretation. A limitation of this study is that 2-weekly sampling does not permit the assessment of hazard on a daily or weekly basis. Moreover, passive sampling does not reflect alterations in the concentrations on short timescales; as such, we are unaware of the extremes in concentration that some analytes might achieve, which will impact our interpretation of the prevalence of antimicrobial resistance genes in this setting. Future research should consider continuous surveillance at a greater number of river sites over multiple seasons, alongside the collection of linked bacterial, population-level and meteorological metadata to better evaluate the impact of ARDCs on driving AMR. Nevertheless, our study identified that in urban Malawian riverine environments, pollution with ARDCs and antibiotics exceeds PNECs considered safe for ecological health over extended periods. This finding is likely to be a consequence of inadequate WASH infrastructure in densely populated urban environments, human antimicrobial usage, and the local climate. Given the local river networks are a point of interaction in daily life, used for bathing, washing clothes, agricultural and animal practices (15), the need for improvements in solid, human, and animal waste management are urgently required to impact the transmission and emergence of AMR, improve the health of residents, and enhance biodiversity. Without improvements to environmental health, we are unlikely to control AMR in these settings.

## Methods

### Site selection, study design and sampling methods

This study was undertaken in Blantyre, southern Malawi, by the Drivers of Resistance in Uganda and Malawi (DRUM) consortium (30). Blantyre has a population of ∼830,000 people, is served by a single 1350-bed tertiary hospital, and has basic citywide sanitation infrastructure, with only one operational wastewater treatment plant (WWTP). An iterative approach to site selection was undertaken during a 9-month pilot phase (February 2020 - October 2020). Initially, 5 sites were identified via transect walks undertaken in the urban and peri-urban districts alongside an area downstream of the city centre (**Figure 1 & Figure S13**). During the pilot phase, river water was purposively collected, and the utility of each site was assessed via a number of logistical and safety parameters (**Table S1**). Given logistical challenges, primarily due to mechanical loss and theft of the passive chemical samplers (**Table S1**), two key rivers were selected for onward sampling (**Figure S14-15**), which represented a river site downstream of the city centre/hospital (site 1) and a river site downstream of a dense urban community (site 2). These 2 sites then underwent uninterrupted sampling over a 12-month period (November 2020 to November 2021).

Polar organic chemical integrative samplers (POCIS) [Nya Exposmeter AB, Trehorningen 34, SE-92266 Tavelsjo, Sweden, www.exposmeter.com] were situated in urban rivers and replaced at 2-weekly intervals. POCIS consists of a sorbent sandwiched between two polyethersulfone membranes, fixed into a porous metal cage (**Figure S16**). The membrane allows for the passage of dissolved chemicals onto the sorbent, where they become sequestered (31). Samplers were placed at 20-100cm depth at the fastest portion of the river and attached via metal wire to a stake on the riverbank, hidden from view. On removal, the sampler cage was detached, and the membrane was washed with deionised water to remove any heavy soiling, before being placed in an aluminium foil bag, sealed, and transported to the laboratory within 2 hrs, whereupon it was stored at -80^°^C. Longitudinal metadata of river water parameters were contemporaneously collected.

In addition, river water sampling for heavy metals was completed at sites 1 & 2 (May 2021 and November 2021). A grab sample of river water was collected in a 30 ml universal container during POCIS recovery. Samples were transported to the local laboratory within 2 hrs and stored at ambient temperature in the dark. Subsequently, water samples were shipped at ambient temperature to the UK Centre for Ecology & Hydrology (United Kingdom) for metal analysis. POCIS filters were transported on dry ice to the University of South Bohemia (Czech Republic) for organic pollutant analysis.

### Chemical and heavy metal analysis

A suite of antimicrobials, medications, insecticides, herbicides, fungicides and metals were analysed based on evidence in the literature for their role in the selection or co-selection of antibiotic resistance genes and to examine *a priori* assumptions about antimicrobial use in Blantyre (**Tables S3a-c**). POCIS samplers were extracted using standard procedures described previously (32). Metals were analysed by inductively coupled plasma mass spectrometry method (Perkin Elmer Nexion 300D ICP-MS). Targeted micropollutants analysis was performed using liquid chromatography coupled with tandem mass spectrometry (TSQ Quantiva mass spectrometer, Accela 1250 pump, both Thermo Fisher Scientific; PAL autosampler, CTC Switzerland)(31,33). Limits of quantification (LOQ) were calculated from the instrumental LOQ by correcting to the internal standard response, for the matrix effect, for internal standard response, and for the aliquot/volume of individual samples (31). Water concentrations of medications were calculated from POCIS adsorbed mass according to field calibration study data (26). The POCIS extracts were analysed using nontargeted LC-high resolution mass spectrometry (QExactive hybrid quadrupole-orbital trap mass spectrometer, Thermo Fisher Scientific) operated in combined full-scan/data independent modes (34). The data were processed using Compound Discoverer 3.1 software to permit the identification of chemical compounds that were present but not included in the targeted analysis, including antiretrovirals, antibiotics, antifungals and antiprotozoals. PCA of nontargeted POCIS extracts data determined site-based differences in chemical compositions.

### Statistical analysis

Statistical analysis and graphic visualisations including ratios, means +/- standard deviation (SD), median +/- interquartile range (IQR), violin plots, Pearson’s coefficient matrix, PCA and PNEC tables were performed using R studio (Version 1.4.11). Sampling site maps were drawn using QGIS (Version 3.4). Analysis of the LOQ for concentrations includes the 1-year continuous dataset without pilot data. Differences in chemical concentrations found at locations and seasons were evaluated via Man-Whitney and signed-rank Wilcoxon tests, respectively. PNEC values were obtained from international guidance and recent literature (**Table S4a-b**) (10). The ratios of monthly analyte mean/PNEC value were used for illustration. CECs were taken for human pharmaceuticals where PNECs do not exist (17,35). The wet season was classified as samples obtained between November and April, and the dry season was classified as samples obtained between May and October.

## Ethical statement

This study was conducted within the DRUM consortium (MRC funded; MR/S004793/1) and as part of a personal fellowship (Wellcome Trust: 216221). The LC-MS analyses were performed at VVI CENAKVA Research Infrastructure (ID 90099, MEYS Czech Republic, 2019–2022), and nontargeted analyses were performed within grant No. 20-04676X funded by the Czech Science Foundation. Ethical approval was obtained for this study from the College of Medicine Research and Ethics Committee, Malawi (P.11/18/2541) and the Liverpool School of Tropical Medicine, UK (18-090). In addition, permissions were granted from village leaders, and support obtained from local community advisory groups. Sensitizations of study areas were conducted prior to study initiation.

## Data availability

All relevant data are included in the manuscript and within the supplementary information.

## Acknowledgements

We thank the local communities for their acceptance of this study and would also like to thank the wider DRUM consortium for their advice, guidance, and support.

## Contributions

DC, AS, and NF conceived the study. DC and AS devised the analysis plan. DC, RG, KG and AVS analysed the data. AS, NF and NE verified and interpreted. DC drafted the initial manuscript. All authors contributed to revision of the manuscript. DC and NF acquired the study funding and DC, ToM, KC, TrM, RG, KG and AVS verified the underlying data. All authors take responsibility for the decision to submit for publication.

## Declaration of interests

We declare no competing interests.

## Additional information

Supplementary information has been included alongside chemical-linked metadata.

## Notes

### Competing Interest Statement

The authors have declared no competing interest.

### Funding Statement

This study was funded by the Medical Research Council, NIHR, Wellcome Trust, the Ministry of Education, Youth and Sports of the Czech Republic, Czech Science Foundation

## References

1. Kimera ZI, Mshana SE, Rweyemamu MM, Mboera LEG, Matee MIN. Antimicrobial use and resistance in food-producing animals and the environment: an African perspective. Antimicrob Resist Infect Control. 2020 Mar 3;9(1):37.

2. Antimicrobial Resistance Collaborators. Global burden of bacterial antimicrobial resistance in 2019: a systematic analysis. Lancet Lond Engl. 2022 Feb 12;399(10325):629–55.

3. O’neill J. Tackling drug-resistant infections globally: Final report and recommendations. Lond HM Gov Wellcome Trust. 2016;

4. Cassini A, Högberg LD, Plachouras D, Quattrocchi A, Hoxha A, Simonsen GS, et al. Attributable deaths and disability-adjusted life-years caused by infections with antibiotic-resistant bacteria in the EU and the European Economic Area in 2015: a population-level modelling analysis. Lancet Infect Dis. 2019 Jan 1;19(1):56–66.

5. Lowder SK, Skoet J, Raney T. The Number, Size, and Distribution of Farms, Smallholder Farms, and Family Farms Worldwide. World Dev. 2016 Nov 1;87:16–29.

6. Larsson DGJ, Flach CF. Antibiotic resistance in the environment. Nat Rev Microbiol. 2022 May;20(5):257–69.

7. Singer AC, Shaw H, Rhodes V, Hart A. Review of Antimicrobial Resistance in the Environment and Its Relevance to Environmental Regulators. Front Microbiol. 2016;7:1728.

8. Singer AC, Xu Q, Keller VDJ. Translating antibiotic prescribing into antibiotic resistance in the environment: A hazard characterisation case study. PLOS ONE. 2019 Sep 4;14(9):e0221568.

9. Polianciuc SI, Gurzău AE, Kiss B, Stefan MG, Loghin F. Antibiotics in the environment: causes and consequences. Med Pharm Rep. 2020 Jul;93(3):231–40.

10. Tell J, Caldwell DJ, Häner A, Hellstern J, Hoeger B, Journel R, et al. Science-based Targets for Antibiotics in Receiving Waters from Pharmaceutical Manufacturing Operations. Integr Environ Assess Manag. 2019;15(3):312–9.

11. Grenni P, Ancona V, Barra Caracciolo A. Ecological effects of antibiotics on natural ecosystems: A review. Microchem J. 2018;136:25–39.

12. Murray LM, Hayes A, Snape J, Kasprzyk-Hordern B, Gaze WH, Murray AK. Co-selection for antibiotic resistance by environmental contaminants. Npj Antimicrob Resist. 2024 Apr 1;2(1):1–13.

13. McEwen SA, Collignon PJ. Antimicrobial Resistance: a One Health Perspective. Microbiol Spectr. 2018 Mar;6(2).

14. Nadimpalli ML, Marks SJ, Montealegre MC, Gilman RH, Pajuelo MJ, Saito M, et al. Urban informal settlements as hotspots of antimicrobial resistance and the need to curb environmental transmission. Nat Microbiol. 2020 Jun;5(6):787–95.

15. Cocker D, Chidziwisano K, Mphasa M, Mwapasa T, Lewis JM, Rowlingson B, et al. Investigating One Health risks for human colonisation with extended spectrum β-lactamase-producing Escherichia coli and Klebsiella pneumoniae in Malawian households: a longitudinal cohort study. Lancet Microbe. 2023 Jul 1;4(7):e534–43.

16. Bengtsson-Palme J, Kristiansson E, Larsson DGJ. Environmental factors influencing the development and spread of antibiotic resistance. FEMS Microbiol Rev. 2018 Jan 1;42(1).

17. Wilkinson JL, Boxall ABA, Kolpin DW, Leung KMY, Lai RWS, Galbán-Malagón C, et al. Pharmaceutical pollution of the world’s rivers. Proc Natl Acad Sci U S A. 2022 Feb 22;119(8):e2113947119.

18. Zhou Q, Yang N, Li Y, Ren B, Ding X, Bian H, et al. Total concentrations and sources of heavy metal pollution in global river and lake water bodies from 1972 to 2017. Glob Ecol Conserv. 2020 Jun 1;22:e00925.

19. Munita JM, Arias CA. Mechanisms of Antibiotic Resistance. Microbiol Spectr. 2016 Apr;4(2): 10.1128/microbiolspec.VMBF-0016–2015.

20. Kumwenda S, Tsakama M, Kalulu K, Kambala C. Determination of Biological, Physical and Chemical Pollutants in Mudi River, Blantyre, Malawi. 2012 Feb 18;2:6833–9.

21. Schreinemachers P, Tipraqsa P. Agricultural pesticides and land use intensification in high, middle and low income countries. Food Policy. 2012 Dec 1;37(6):616–26.

22. Murray AK, Stanton IC, Tipper HJ, Wilkinson H, Schmidt W, Hart A, et al. A critical meta-analysis of predicted no effect concentrations for antimicrobial resistance selection in the environment. Water Res. 2024 Nov 15;266:122310.

23. Faleye AC, Adegoke AA, Ramluckan K, Bux F, Stenström TA. Antibiotic Residue in the Aquatic Environment: Status in Africa. Open Chem. 2018 Jan 1;16(1):890–903.

24. MacPherson EE, Reynolds J, Sanudi E, Nkaombe A, Mankhomwa J, Dixon J, et al. Understanding antimicrobial use in subsistence farmers in Chikwawa District Malawi, implications for public awareness campaigns. PLOS Glob Public Health. 2022 Jun 8;2(6):e0000314.

25. Dixon J, MacPherson EE, Nayiga S, Manyau S, Nabirye C, Kayendeke M, et al. Antibiotic stories: a mixed-methods, multi-country analysis of household antibiotic use in Malawi, Uganda and Zimbabwe. BMJ Glob Health. 2021 Nov 1;6(11):e006920.

26. Musicha P, Morse T, Cocker D, Mugisha L, Jewell CP, Feasey NA. Time to define One Health approaches to tackling antimicrobial resistance. Nat Commun. 2024 Oct 10;15(1):8782.

27. Mwapasa T, Chidziwisano K, Mphasa M, Cocker D, Rimella L, Amos S, et al. Key environmental exposure pathways to antimicrobial resistant bacteria in southern Malawi: A SaniPath approach. Sci Total Environ. 2024 Oct 1;945:174142.

28. Chidziwisano K, Cocker D, Mwapasa T, Amos S, Feasey NA, et al. Risk Perception and Psychosocial Factors Influencing Exposure to Antimicrobial Resistance through Environmental Pathways in Malawi. Am J Trop Med Hyg. 2024 Mar 12

29. Musicha P, Beale MA, Cocker D, Oruru FA, Zuza A, Salifu C, et al. One Health in Eastern Africa: No barriers for ESBL producing E. coli transmission or independent antimicrobial resistance gene flow across ecological compartments. bioRxiv. 2024 Jan 1;2024.09.18.613694.

30. Cocker D, Sammarro M, Chidziwisano K, Elviss N, Jacob ST, Kajumbula H, et al. Drivers of Resistance in Uganda and Malawi (DRUM): a protocol for the evaluation of One-Health drivers of Extended Spectrum Beta Lactamase (ESBL) resistance in Low-Middle Income Countries (LMICs). Wellcome Open Res. 2022;7:55.

31. Vrana B, Urík J, Fedorova G, Švecová H, Grabicová K, Golovko O, et al. In situ calibration of polar organic chemical integrative sampler (POCIS) for monitoring of pharmaceuticals in surface waters. Environ Pollut. 2021 Jan 15;269:116121.

32. Fedorova G, Golovko O, Randak T, Grabic R. Passive sampling of perfluorinated acids and sulfonates using polar organic chemical integrative samplers. Environ Sci Pollut Res Int. 2013 Mar;20(3):1344–51.

33. Grabic R, Fick J, Lindberg RH, Fedorova G, Tysklind M. Multi-residue method for trace level determination of pharmaceuticals in environmental samples using liquid chromatography coupled to triple quadrupole mass spectrometry. Talanta. 2012 Oct 15;100:183–95.

34. Nováková P, Švecová H, Bořík A, Grabic R. Novel nontarget LC-HRMS-based approaches for evaluation of drinking water treatment. Environ Monit Assess. 2023 May 26;195(6):739.

35. Fick J, Lindberg RH, Tysklind M, Larsson DGJ. Predicted critical environmental concentrations for 500 pharmaceuticals. Regul Toxicol Pharmacol. 2010 Dec 1;58(3):516–23.

