## Supplemental data for "Environmental hazards from pollution of antibiotics and resistance-driving chemicals in urban river networks of Malawi"

### SUPPLIMENTARY INFORMATION

#### LIST OF TABLES AND FIGURES


*Table S1. Strategy and logistical challenges at sampler sites.* 3

*Table S2. Presence of heavy metals and mean (SD) concentrations (µg/L).* 4

*Table S3a. List of antibiotics and their metabolites screened in river water samples.* 5

*Table S3b. List of insecticides, pesticides, herbicides, fungicides, industrial chemicals, recreational and common human use pharmaceuticals screened in river water samples.* 7

*Table S3c. List of metals screened in river water samples.* 8

*Table S4a. List of PNEC values (ng/L) adapted from the AMR industrial alliance discharge targets.* 9

*Table S4b. List of PNEC and CEC values (ng/L) used in Wilkinson JL et al* 11

*Figure S1. Non-targeted chemical analysis results. (a) PCA analysis of chemical compounds in site 1 and site 2, (b) concentration of novel antibiotics identified through non-targeted method, (c) concentration of ARVs, antiprotozoals and antifungals identified at all sites including those involved in the pilot phase.* 12

*Figure S2a. Presence and absence of insectidices and pesticides over a 1-year period at Site 1.* 13

*Figure S2b. Presence and absence of insecticides and pesticides over a 1-year period at Site 2.* 13

*Figure S2c. Presence and absence of herbicides over a 1-year period at Site 1.* 14

*Figure S2d. Presence and absence of herbicides over a 1-year period at Site 2.* 15

*Figure S2e. Presence and absence of fungicides over a 1-year period at Site 1.* 16

*Figure S2f. Presence and absence of fungicides over a 1-year period at Site 2.* 16

*Figure S2g. Presence and absence of medications over a 1-year period at Site 1, stratified by medication category (i.e. CNS, cardiovascular)* 17

*Figure S2h. Presence and absence of medications over a 1-year period at Site 2, stratified by medication category (i.e. CNS, cardiovascular)* 18

*Figure S2i. Presence and absence of antibiotics over a 1-year period at Site 1, stratified by antibiotic class.* 19

*Figure S2j. Presence and absence of antibiotics over a 1-year period at Site 2, stratified by antibiotic class.* 19

*Figure S3a. Spatiotemporal variations in the chemical compounds found at Site 1 over a 1-year period, presented as the percentage (%) of the total sample concentration (ng/POCIS$^{-1}$/day$^{-1}$) representing each chemical class.* 20

*Figure S3b. Spatiotemporal variations in the chemical compounds found at Site 2 over a 1-year period, presented as the percentage (%) of the total sample concentration (ng/POCIS$^{-1}$/day$^{-1}$) representing each chemical class.* 20

*Figure S4a . Spatiotemporal variations in medication compositions at SITE 1, presented as the percentage (%) of the total medication concentration (ng/POCIS$^{-1}$/day$^{-1}$), representing each medication.* 21

*Figure S4b . Spatiotemporal variations in medication compositions at SITE 2, presented as the percentage (%) of the total medication concentration (ng/POCIS$^{-1}$/day$^{-1}$), representing each medication.* 21

*Figure S5a. Spatiotemporal variations in antibiotic compositions at SITE 1, coloured by antibiotic class.* 22

*Figure S5b. Spatiotemporal variations in antibiotic compositions at SITE 2, coloured by antibiotic class.* 22

*Figure S6a. Violin plot of insecticide and pesticide concentrations (ng/POCIS$^{-1}$/day$^{-1}$) obtained from urban sites.* 23

*Figure S6b. Violin plot of herbicide concentrations (ng/POCIS$^{-1}$/day$^{-1}$) obtained from urban sites.* 23

*Figure S6c. Violin plot of fungicide concentrations (ng/POCIS$^{-1}$/day$^{-1}$) obtained from urban sites.* 24

*Figure S6d. Violin plot of industrial chemical concentrations (ng/POCIS$^{-1}$/day$^{-1}$) obtained from urban sites.* *24*

*Figure S6e. Violin plot of recreational drug and analgesic medication concentrations (ng/POCIS$^{-1}$/day$^{-1}$) obtained from urban sites.* *25*

*Figure S6f. Violin plot ofcommon human-use pharmecutical concentrations (ng/POCIS$^{-1}$/day$^{-1}$) obtained from urban sites.* *25*

*Figure S6g. Violin plot of antibiotic concentrations (ng/POCIS$^{-1}$/day$^{-1}$) obtained from urban sites.* *26*

*Figure S7a. Cumulative total of medications identified from SITE 1, stratified by site and coloured by medication class.* *26*

*Figure 7b. Cumulative total of medications identified from SITE 2, stratified by site and coloured by medication class.* *27*

*Figure S8a. . Temporal relationships in the recovery and concentrations of medications in river water from SITE 1 stratified into safe and unsafe PNEC / CEC levels* *27*

*Figure S8b. Temporal relationships in the recovery and concentrations of medications in river water from SITE 2 stratified into safe and unsafe PNEC / CEC levels.* *28*

*Figure S9. Cumulative total of chemical compounds identified, stratified by site and coloured by chemical class.* *28*

*Figure S10a. Cumulative total of antibiotics (ng/POCIS$^{-1}$/day$^{-1}$) identified from SITE 1, stratified by site and coloured by antibiotic class (Wet season = blue background, Dry season = white background).* *29*

*Figure S10b. Cumulative total of antibiotics (ng/POCIS$^{-1}$/day$^{-1}$) identified from SITE 2, stratified by site and coloured by antibiotic class (Wet season = blue background, Dry season = white background).* *30*

*Figure S11. Pearson's matrix of antibiotics in river water across all 5 study sites. Correlation coefficients are illustrated on a colour spectrum, with those in red and orange showing the highest degree of relationship.* *30*

*Figure S12a. Temporal relationships in the recovery and concentrations of cumulative macrolide risk in river water, stratified by safe and unsafe PNEC levels for each site.* *31*

*Figure S12b. Temporal relationships in the recovery and concentrations of cumulative fluoroquinolone risk in river water, stratified by safe and unsafe PNEC levels for each site.* *31*

*Figure S13. Detailed maps of the riverine network of Blantyre, including (a) Blantyre city (b) Ndirande and (c) Chileka. DRUM study polygons have been demarcated in orange. Sampling sites have been geolocated (site 1: star, site 2: triangle, site 3: square, site 4: circle, site 5: diamond) alongside the key rivers (black = Mudi river, red = Nasolo river, blue = unnamed river).* *32*

*Figure S14. Photos of the sampling sites at pilot study initiation. Local approvals and permissions were granted.* *33*

*Figure S15. Seasonal changes in the rivers at sampling sites. Photos were taken during the pilot and continuous phase, after approvals and local permissions were granted.* *34*

*Figure S16. Porous metal cage (a) sandwiches the PES membrane (b) allowing for environmental exposure while protecting the membrane integrity, which is attached to a metal wire that is secured to the river bank (c).* *34*

**Table S1. STRATERGY AND LOGISTICAL CHALLENGES AT SAMPLER SITES**

Local chiefs and community leaders were surveyed for the acceptance of samplers and verbal permissions were granted. Where sites fell on private property, verbal agreements were drafted for placement of samplers prior to siting.

A pilot phase between February 2020 and October 2020 was conducted to assess for logistical challenges at 5 sites in Blantyre, Malawi. Three of the sites faced significant challenges from theft and the unsuccessful recovery of filters after either a 7 day or 14-day period. In the furthest downstream river site, high fluctuations in rainfall during the "rainy" season led to flash flooding and mechanical destruction of samplers, while 2 sites situated in dense urban environments where the samplers were clearly visible, had high levels of theft.

Given the study required continuous sampling over a 1-year period to enable longitudinal assessment of antibiotic presence to evaluate seasonal effects, in November 2020 a rational approach to sampling was undertaken for the study, with a focus on 2 sites in urban riverine environments alone (**Figure 1 and S1**).

Additionally, advice was sought from local community research groups and leaders as to the best approaches to deal with some of the issues identified during the pilot phase. In line with these conversations, some mechanical and technical alterations to the recovery and placement of the filters were made. The samplers were placed into a small surrounding cage to reduce the risk of mechanical destruction (pictured in Figure S4, adapted from "Instillation of POCIS samplers" by R Grabic, University of South Bohemia), and then submerged into the river. Samplers were attached via a wire and secondary rope to metal posts drilled into the edge of the riverbank at points out of view from the public. The length of wire was ~20cm long to enable continuous submersal and limited movement. Collection and replacement of samplers were undertaken at times of reduced footfall, by members of the community known to the study team, in an effort to reduce the chance of filter discovery and theft.

**Table S2. PRESENCE OF HEAVY METALS AND MEDIAN (IQR) CONCENTRATIONS (μg/L).**

| Metal | Site 1 median (IQR) | Site 2 median (IQR) | Metal | Site 1 median (IQR) | Site 2 median (IQR) |
|---|---|---|---|---|---|
| Al | 11.4 (35.71) | 10.72 (32.41) | Mo | 1.05 (0.36) | 0.95 (0.47) |
| As | 0.68 (0.19) | 1.35 (0.64) | Ni | 11.7 (6.22) | 0.4 (0.42) |
| Ba | 116 (52.6) | 139.5 (72.95) | Pb | 0.01 (0.02) | < LOQ |
| Be | < LOQ | < LOQ | Rb | 12.3 (4.35) | 39.05 (19.82) |
| Cd | 0.01 (0.01) | < LOQ | Sb | 17.7 (15.23) | 0.53 (0.19) |
| Ce | 0.01 (0.01) | 0.01 (0.01) | Se | 0.43 (0.19) | 0.56 (0.16) |
| Co | 0.29 (0.34) | 0.23 (0.57) | Sn | < LOQ | < LOQ |
| Cr | 4.65 (2.53) | 0.66 (0.37) | Sr | 511 (73) | 952 (291) |
| Cs | 0.01 (0.01) | 0.04 (0.01) | Ti | 7.24 (11.02) | 8.64 (8.39) |
| Cu | 6.17 (3.91) | 4.32 (1.02) | U | 0.1 (0.06) | 0.03 (0.03) |
| Fe | 22.5 (30.95) | 9.88 (5.25) | V | 2.38 (1.61) | 1.7 (1.48) |
| La | 0.01 (0.00) | < LOQ | W | 0.01 (0.04) | < LOQ |
| Li | 1.09 (0.21) | 3.17 (0.53) | Zn | 25.42 (34.09) | 5.56 (3.25) |
| Mn | 2.23 (6.09) | 1.88 (6.42) | | | |

**Table S3a. LIST OF ANTIBIOTICS AND THEIR METABOLITES SCREENED IN RIVER WATER SAMPLES**

| Antibiotic Class | Antibiotic Name | Acronym |
|---|---|---|
| β-lactams<br>(β-Ls) | Amoxicillin<br>Ampicillin<br>Cloxacillin<br>Flucloxacillin<br>Penicillin G<br>Penicillin V | AMX<br>AMP<br>CLX<br>FLX<br>PENG<br>PENV |
| | Cefalexin<br>Cefixime<br>Cefotaxime<br>Cefuroxime<br>Ceftriaxone | CEF<br>CFX<br>CTX<br>CXM<br>CRO |
| Quinolones (QNs) | Ciprofloxacin<br>Difloxacin<br>Enoxacin<br>Enrofloxacin<br>Flumequine<br>Levofloxacin + Ofloxacin<br>Lomefloxacin<br>Norfloxacin<br>Oxolinic acid<br>Perfloxacin<br>Roxithromycin | CIP<br>DIF<br>ENX<br>EFX<br>FLU<br>LEV<br>LOM<br>NOR<br>OXO<br>PER<br>ROX |
| MLS drugs<br>*(MLS)* | Azithromycin<br>Clarithromycin<br>Clindamycin<br>Clindamycin sulfoxide<br>Erythromycin<br>Tylosin | AZM<br>CLR<br>CLI<br>CLS<br>ERY<br>TYL |
| Sulphonamides<br>*(SAs)* | Sulfadiazine<br>Sulfamerazine<br>Sulfamethazine<br>Sulfamethizole<br>Sulfamethoxazole<br>Sulfamethoxine<br>Sulfamethoxypyridine<br>Sulfamoxole<br>Sulfaphenazole<br>Sulfapyridine<br>Sulfaquinoxaline<br>N1 Acetyl SMX<br>N4 Acetyl SMX<br>Sulfathiazole | SFD<br>SFM<br>SFT<br>SFZ<br>SMX<br>SMI<br>SMP<br>SML<br>SPZ<br>SPY<br>SFQ<br>NA1<br>NA4<br>STZ |
| Tetracyclines<br>*(TCs)* | Chlortetracycline<br>Doxycycline<br>Oxytetracycline<br>Tetracycline | CLT<br>DOX<br>OXY<br>TET |

| | | |
|---|---|---|
| Other antibiotics<br>*(Other)* | Chloramphenicol<br>Florfenicol<br>Metronidazole<br>Rifampicin<br>Trimethoprim | CHL<br>FLO<br>MET<br>RIF<br>TRI |
| Antifungals and Antiprotozoals<br>*(Fung)* | Ornidazole | ORN |
| | Miconazole<br>Terbinafine<br>Clotrimazole<br>Ketoconazole | MIC<br>TER<br>CLZ<br>KET |

**Table S3b. LIST OF INSECTICIDES, HERBICIDES, FUNGICIDES, INDUSTRIAL CHEMICALS AND HUMAN USE PHARMACEUTICALS SCREENED IN RIVER WATER SAMPLES**

| Class | Chemicals and metabolites |
|---|---|
| **Insecticides and metabolites** | Carbofuran-3-hydroxy, Chlorantraniliprole, Chlorpyrifos, DEET, Diazinon, Dimethoate, Imidacloprid, Malathion,Methoxyfenozide, Pirimicarb, Pirimiphos_ethyl, Pirimiphos_methyl, Thiamethoxam, Warfarin |
| **Herbicides and metabolites** | 1-(3,4-Dichlorophenyl)_urea, 2,4,5-Trichlorophenoxyacetic_acid, 2,4-Dichlorophenoxyacetic_acid, 2,4-Dichlorophenoxypropionic_acid, 3-chloro-4-methylaniline, 4-Isopropylaniline, Acetochlor, Acetochlor_ESA, Acetochlor_OA, Alachlor, Alachlor_ESA, Alachlor_OA, Ametryn, Anthranilic_acid_isopropylamide, Atraton,Atrazine, Atrazine_2-hydroxy, Atrazine_desethyl, Atrazine_desethyl-2-hydroxy, Atrazine_desethyl-desisopropyl, Atrazine_desisopropyl, Bensulfuron_methyl, Bentazone, Chloridazon, Chloridazon_desphenyl, Chloridazon_methyl_desphenyl, Chlorotoluron, Chlorotoluron_desmethyl, Clomazone, Cyanazine, Desmetryn, Dimethachlor, Dimethachlor_ESA, Dimethachlor_OA, Dimethenamid_ESA, Dimethenamid_OA, Diuron, Diuron_desmethyl, Fenuron, Florasulam, Fluazifop-p, Foramsulfuron, Hexazinone, Imazamethabenz_methyl, Imazamox, Ioxynil, Isoproturon, Isoproturon_didemethyl, Isoproturon_monodemethyl, Lenacil, Linuron, MCPA, MCPP, Metazachlor, Metazachlor_ESA, Metazachlor_OA, Methabenzthiazuron, Metobromuron, Metolachlor, Metolachlor_ESA, Metolachlor_OA, Metoxuron, Metribuzin, Metribuzin_desamino, Metsulfuron_methyl, Monolinuron, N-chloroacetyl-2,6-diethylaniline, Picloram, Prometryn, Propachlor, Propazine, Propazine_hydroxy, Sebuthylazine, Simazine, Simazine_hydroxy, Terbuthylazine, Terbuthylazine_desethyl, Terbuthylazine_desethyl-2-hydroxy, Terbuthylazine_hydroxy, Terbutryn,, Triallate |
| **Fungicides** | Azoxystrobin, Carbendazim, Cyproconazole, Dimethomorph, Epoxiconazole, Flusilazole, Metalaxyl, Metconazole, Propiconazole, Pyrimethanil, Tebuconazole, Triadimenol, Triticonazole |
| **Persistent industrial chemicals** | 1H-benzotriazol, 1H-benzotriazol_(5/4)-methyl, 1H-benzotriazol_1-methyl |
| **Recreational and analgesic pharmaceuticals** | 2-oxo-3-hydroxy-LSD, 6-acetylmophine, Amphetamine, Benzoylecgonine, Cannabinol, Catinone, Cocaine, Ketamine, MDA, MDEA, MDMA, Mephedrone, Metamphetamine, Methadone, Morphine, Norketamine, Oxycodone, THC-COOH |
| **Common human-use pharmaceuticals** | Alfuzosin, Alprazolam, Amitryptyline, Atenolol, Atorvastatin, Bezafibrate, Biperiden, Bisoprolol, Caffeine, Carbamazepine (CBZ), Dihydro CBZ, Epoxy CBZ, trans-dihydro-dihydroxy CBZ, Cetirizine, Cilazapril, Citalopram, N-desmethylcitalopram, Clemastine, Clomipramine, Clonazepam, Codeine, Diclofenac, Dicycloverine, Diltiazem, Diphenhydramine, Disopyramide, Donepezil, Eprosartan, Fenofibrate, Fexofenadine, Gabapentin, Glibenclamide, Glimepiride, Haloperidol, Iopromide, Irbesartan, Lamotrigine, Loperamide, Maprotiline, Meclizine, Memantine, Metoprolol, Metoprolol acid, Mianserin, Mirtazapine, Orphenadrine, Oseltamivir carboxylate, Oxazepam, Oxcarbazepine, Paroxetine, Pizotifen, Propranolol, Ropinirole, Rosuvastatin, Sertraline, Norsertraline, Sotalol, Sulfasalazine, Tamoxifen, Telmisartan, Terbutaline, Theophylline, Tramadol (TRM), N-desmethylTRM, O-desmethylTRM, Trazodone, Triamterene, Valsartan, Venlafaxine, O-Desmethylvenlafaxine, Verapamil, Vortioxetine |

**Table S3c. LIST OF METALS SCREENED IN RIVER WATER SAMPLES**

| Metal | Acronym | Metal | Acronym |
|---|---|---|---|
| Aluminium | Al | Molybdenum | Mo |
| Arsenic | As | Nickel | Ni |
| Barium | Ba | Lead | Pb |
| Beryllium | Be | Rubidium | Rb |
| Cadmium | Cd | Antimony | Sb |
| Cerium | Ce | Selenium | Se |
| Cobalt | Co | Tin | Sn |
| Chromium | Cr | Strontium | Sr |
| Caesium | Cs | Titanium | Ti |
| Copper | Cu | Uranium | U |
| Iron | Fe | Vanadium | V |
| Lanthanum | La | Tungsten | W |
| Lithium | Li | Zinc | Zn |
| Manganese | Mn | | |

**Table S4a. LIST OF PNEC VALUES (ng/L) ADAPTED FROM THE AMR INDUSTRIAL ALLIANCE DISCHARGE TARGETS**

| Antibiotic | PNEC | Antibiotic | PNEC | Antibiotic | PNEC |
|---|---|---|---|---|---|
| Amikacin | 16 | Cloxacillin | 0.13 | Oxytetracycline | 0.5 |
| Amoxicillin | 0.25 | Colistin | 2.0 | Pefloxacin | 8.0 |
| Amphotericin B | 0.02 | Daptomycin | 1.0 | Phenoxymethylpenicillin | 0.06 |
| Ampicillin | 0.25 | Delamanid | 0.03 | Piperacillin | 0.5 |
| Anidulafungin | 0.02 | Doripenem | 0.11 | Polymixin B | 0.06 |
| Avibactam | 200 | Doxycycline | 2.0 | Retapamulin | 0.06 |
| Avilamycin | 8.0 | Enramycin | 4.8 | Rifampicin | 0.06 |
| Azithromycin | 0.02 | Enrofloxacin | 0.06 | Roxithromycin | 1.0 |
| Aztreonam | 0.5 | Ertapenem | 0.13 | Secnidazole | 1.0 |
| Bacitracin | 8.0 | Erythromycin | 0.5 | Sparfloxacin | 0.06 |
| Bedaquiline | 0.08 | Ethambutol | 2.0 | Spectinomycin | 32 |
| Benzylpenicillin | 0.25 | Faropenem | 0.02 | Spiramycin | 0.5 |
| Capreomycin | 2.0 | Fidaxomicin | 0.02 | Streptomycin | 16 |
| Cefaclor | 0.50 | Florfenicol | 2.0 | Sulbactam | 16 |
| Cefadroxil | 2.0 | Fluconazole | 0.25 | Sulfadiazine | 13 |
| Cefalonium | 21 | Flumequine | 0.25 | Sulfamethoxazole | 0.6 |
| Cefaloridine | 4.0 | Fosfomycin | 2.0 | Tedizolid | 3.2 |
| Cefalothin | 2.0 | Fusidic acid | 0.5 | Teicoplanin | 0.5 |
| Cefazolin | 1.0 | Gatifloxacin | 0.13 | Telithromycin | 0.06 |
| Cefdinir | 0.25 | Gemifloxacin | 0.06 | Tetracycline | 1.0 |
| Cefepime | 0.5 | Gentamicin | 0.15 | Thiamphenicol | 1.0 |
| Cefixime | 0.06 | Imipenem | 0.13 | Tiamulin | 1.0 |
| Cefoperazone | 0.5 | Isoniazid | 0.13 | Ticarcillin | 8.0 |
| Cefotaxime | 0.1 | Itraconazole | 0.01 | Tigecycline | 1.0 |
| Cefoxitin | 8.0 | Kanamycin | 1.0 | Tildipirosin | 0.42 |
| Cefpirome | 0.06 | Levofloxacin | 0.25 | Tilmicosin | 1.0 |
| Cefpodoxime | 0.25 | Lincomycin | 0.81 | Tobramycin | 1.0 |

| | | | | | |
|---|---|---|---|---|---|
| Cefquinome | 1.6 | Linezolid | 6.7 | Trimethoprim | 0.5 |
| Ceftaroline | 0.06 | Loracarbef | 2.0 | Trovafloxacin | 0.03 |
| Ceftazidime | 0.5 | Mecillinam | 1.0 | Tylosin | 1.0 |
| Ceftibuten | 0.25 | Meropenem | 0.06 | Vancomycin | 8.0 |
| Ceftiofur | 0.06 | Metronidazole | 0.13 | Viomycin | 2.0 |
| Ceftobiprole | 0.23 | Minocycline | 1.0 | Virginiamycin | 2.0 |
| Ceftolozane | 1.9 | Moxifloxacin | 0.13 | | |
| Ceftriaxone | 0.03 | Mupirocin | 0.25 | | |
| Cefuroxime | 0.5 | Nalidixic acid | 16 | | |
| Cephalexin | 0.08 | Narasin | 0.5 | | |
| Cephradine | N/A | Neomycin | 0.03 | | |
| Chloramphenicol | 8.0 | Netilmicin | 0.5 | | |
| Ciprofloxacin | 0.06 | Nitrofurantoin | 64 | | |
| Clarithromycin | 0.08 | Norfloxacin | 0.5 | | |
| Clinafloxacin | 0.5 | Ofloxacin | 0.5 | | |
| Clindamycin | 0.1 | Oxacillin | 1.0 | | |

**Table S4b. LIST OF PNEC AND CEC VALUES (ng/L) USED IN WILKINSON JL ET AL.**

| **Medication** | **PNEC/CEC*** | **Medication** | **PNEC/CEC*** | **Medication** | **PNEC/CEC*** |
|---|---|---|---|---|---|
| Amitriptyline* | 48 | Fexofenadine* | 20222 | Oxazepam* | 30721 |
| Amoxicillin | 250 | Fluconazole | 25 | Oxytetracycline | 500 |
| Atenolol | 148000 | Fluoxetine* | 489 | Paracetamol* | 24000000 |
| Carbamazepine | 25000 | Gabapentin | 450000000 | Pregabalin | 100000 |
| Cetirizine* | 423061 | Itraconazole | 8 | Propranolol | 20 |
| Cimetidine | 176000 | Ketoconazole | 50 | Ranitidine* | 232954 |
| Ciprofloxacin | 60 | Ketotifen* | 12 | Salbutamol* | 27669 |
| Citalopram* | 141 | Lidocaine* | 466820 | Sertraline* | 51 |
| Clarithromycin | 250 | Lincomycin | 810 | Sitagliptin | 390000 |
| Cloxacillin | 130 | Loratadine | 0.56 | Sulfadiazine | 11210 |
| Codeine* | 26620 | Metformin | 1000000 | Sulfamethoxazole | 200 |
| Diazepam | 7800 | Metronidazole | 130 | Tetracycline | 1000 |
| Diltiazem* | 27884 | Miconazole | 200 | Tramadol* | 4799 |
| Diphenhydramine* | 2035 | Naproxen | 150000 | Trimethoprim | 500 |
| Enrofloxacin | 60 | Nevirapine | 12070 | Venlafaxine* | 6112 |
| Erythromycin | 500 | Norethisterone* | 486 | Verapamil* | 24 |

*Indicates predicted Critical Environmental Concentrations (CECs). CEC values have been obtained from those used in Wilkinson JL et al, which were originally predicted to represent the surface water concentrations in pharmaceuticals that would be expected to cause a pharmacological effect in fish.

**Figure S1**. **NON-TARGETTED CHEMICAL ANALYSIS RESULTS.** (a) PCA analysis of chemical compounds in site 1 and site 2, (b) presence of antiretrovirals (ARV), antimalarials (AM) and anti-tuberculous medications (ATB) identified through non-targeted chemical analysis.

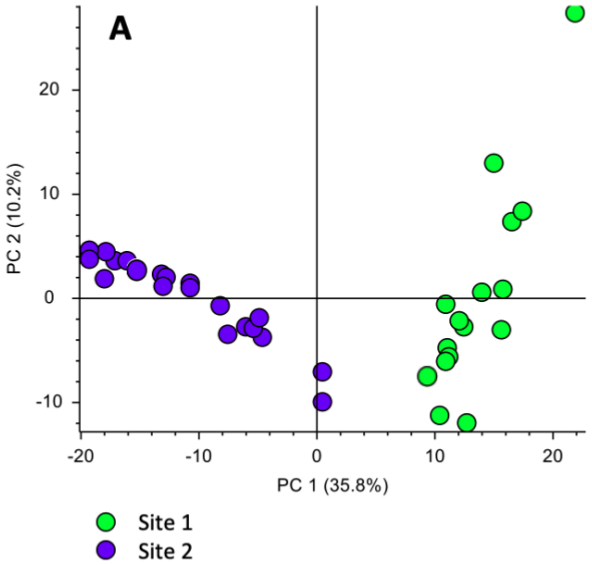


**B**

| Medication | Class | CAS No | Molecular formula | RT | Annot. DeltaMass [ppm] | Library used | Schmymanski level§ | Dry season | | Wet season | |
|---|---|---|---|---|---|---|---|---|---|---|---|
| | | | | | | | | Site 1 | Site 2 | Site 1 | Site 2 |
| **Abacavir** | **ARV** | 136470-78-5 | C14 H18 N6 O | 6.04 | 1.24 | mzCloud | 2 | 95% | 97% | 93% | 97% |
| **Nevirapine** | **ARV** | 129618-40-2 | C15 H14 N4 O | 7.97 | 0.74 | mzCloud | 2 | 100% | 100% | 100% | 97% |
| **Lamivudine** | **ARV** | 131086-21-0 | C8 H11 N3 O3 S | 1.12 | -0.87 | massBanku EU | 3 | 9% | 0% | 66% | 93% |
| **Efavirenz** | **ARV** | 154598-52-4 | C14 H9 Cl F3 N O2 | 10.50 | 0.97 | mzCloud | 2 | 91% | 93% | 45% | 97% |
| **Zidovudine** | **ARV** | 30516-87-1 | C10 H13 N5 O4 | 1.66 | 0.39 | massBanku EU | 3 | 27% | 3% | 69% | 23% |
| **Lopinavir** | **ARV** | 192725-17-0 | C37 H48 N4 O5 | 10.82 | 0.86 | mzCloud | 2 | 95% | 97% | 97% | 83% |
| **Sulfadoxine** | **AM** | 2447-57-6 | C12 H14 N4 O4 S | 6.64 | 0.43 | mzCloud | 2 | 91% | 97% | 97% | 97% |
| **Atazanavir** | **ARV** | 198904-31-3 | C38 H52 N6 O7 | 10.63 | 0.42 | mzCloud | 2 | 95% | 97% | 97% | 87% |
| **Ritonavir** | **ARV** | 155213-67-5 | C37 H48 N6 O5 S2 | 10.66 | 1 | mzCloud | 2 | 73% | 83% | 17% | 93% |
| **Pyrimethamine** | **AM** | 58-14-0 | C12 H13 Cl N4 | 7.48 | 1.49 | massBanku EU | 3 | 77% | 69% | 79% | 23% |
| **Isoniazid** | **ATB** | 54-85-3 | C6 H7 N3 O | 5.42 | 1.82 | mzCloud | 2 | 32% | 14% | 10% | 0% |

§ … Schymanski level according to a scale published in: Schymanski, E.L., Jeon, J., Gulde, R., Fenner, K., Ruff, M., Singer, H.P., Hollender, J., 2014. Identifying Small Molecules via High Resolution Mass Spectrometry: Communicating Confidence. Environmental Science & Technology 48, 2097-2098.

**Figure S2a. PRESENCE AND ABSENCE OF INSECTICIDES OVER A 1-YEAR PERIOD AT SITE 1.**

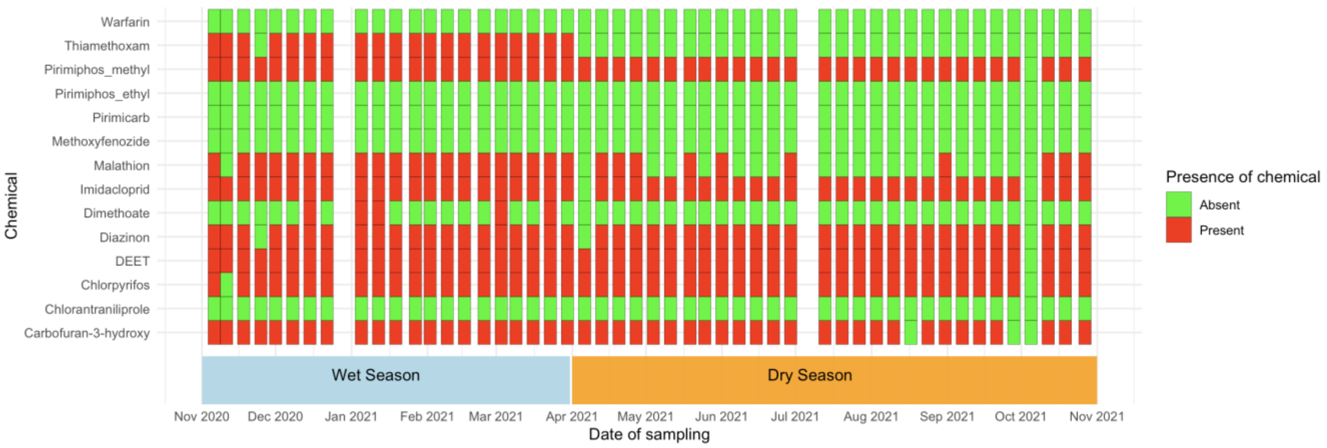


**Figure S2b. PRESENCE AND ABSENCE OF INSECTICIDES OVER A 1-YEAR PERIOD AT SITE 2**

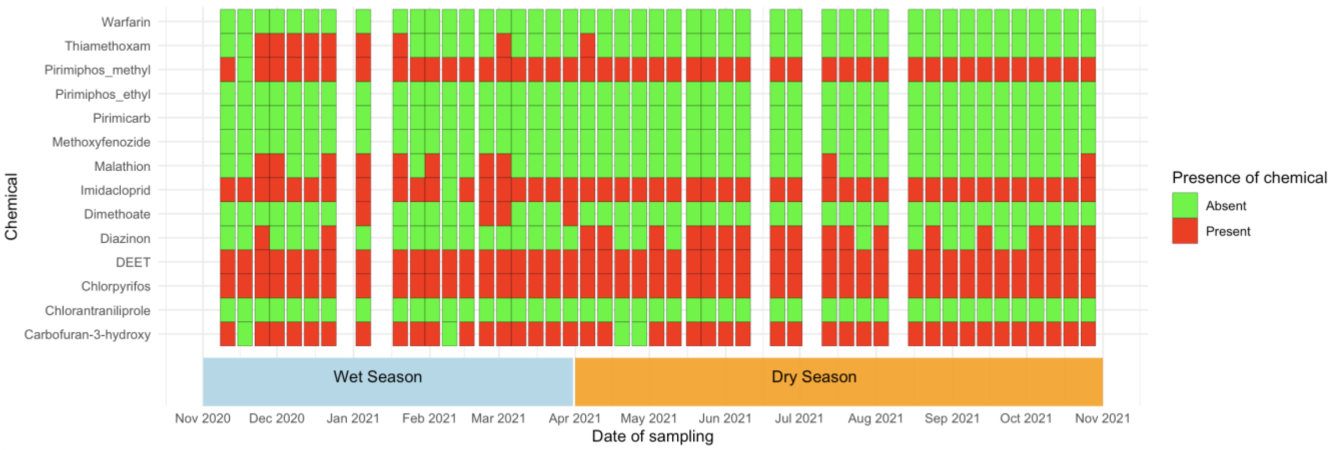

**Figure S2c. PRESENCE AND ABSENCE OF HERBICIDES OVER A 1-YEAR PERIOD AT SITE 1**

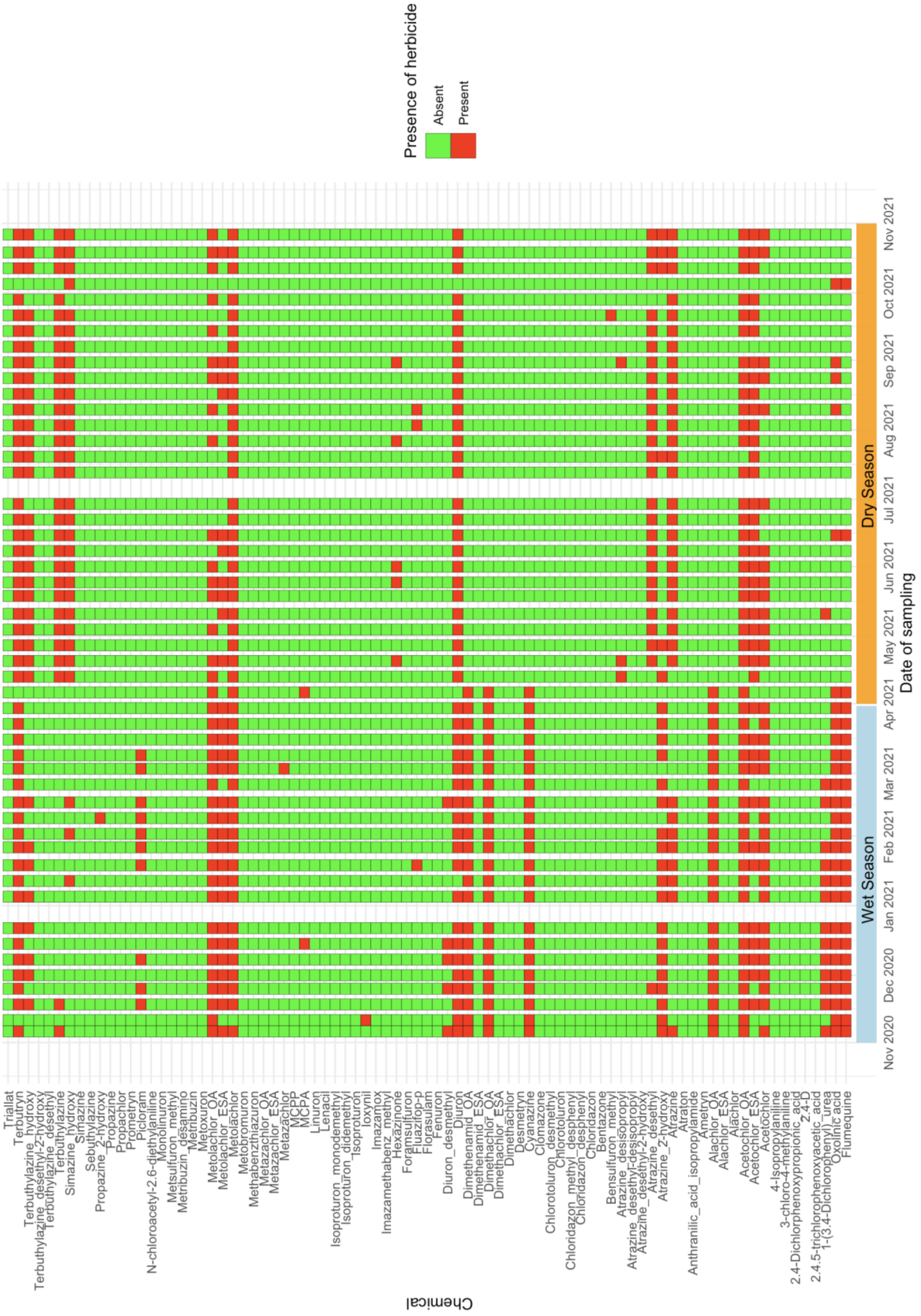

**Figure S2d. PRESENCE AND ABSENCE OF HERBICIDES OVER A 1-YEAR PERIOD AT SITE 2**

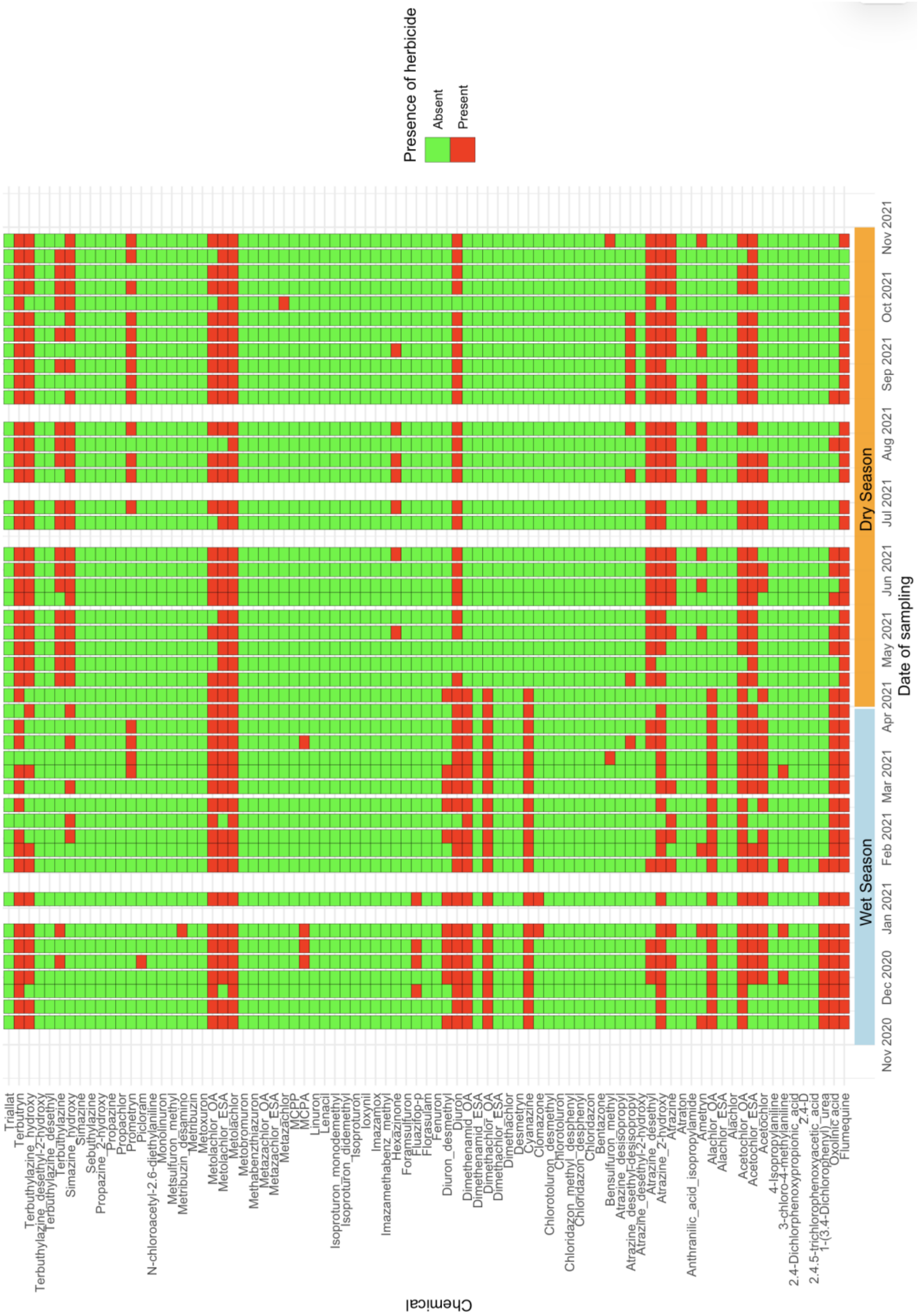

**Figure S2e. PRESENCE AND ABSENCE OF FUNGICIDES OVER A 1-YEAR PERIOD AT SITE 1.**

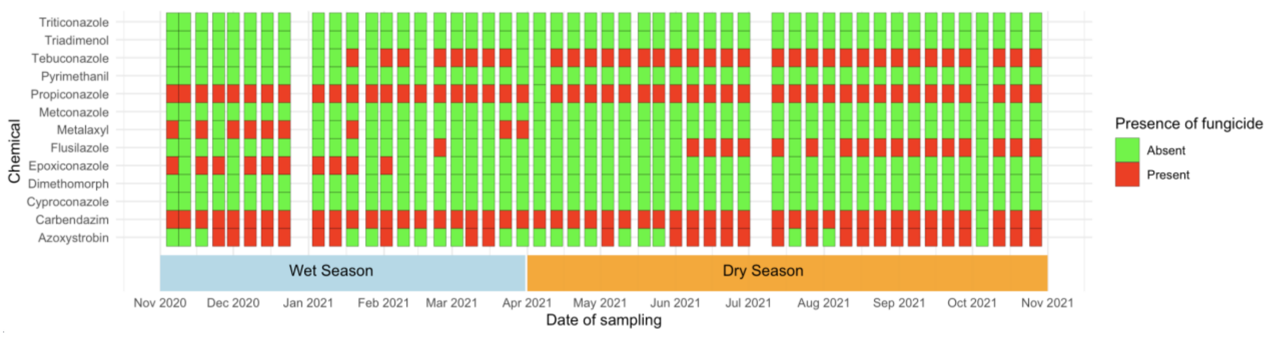


**Figure S2f. PRESENCE AND ABSENCE OF FUNGICIDES OVER A 1-YEAR PERIOD AT SITE 2**

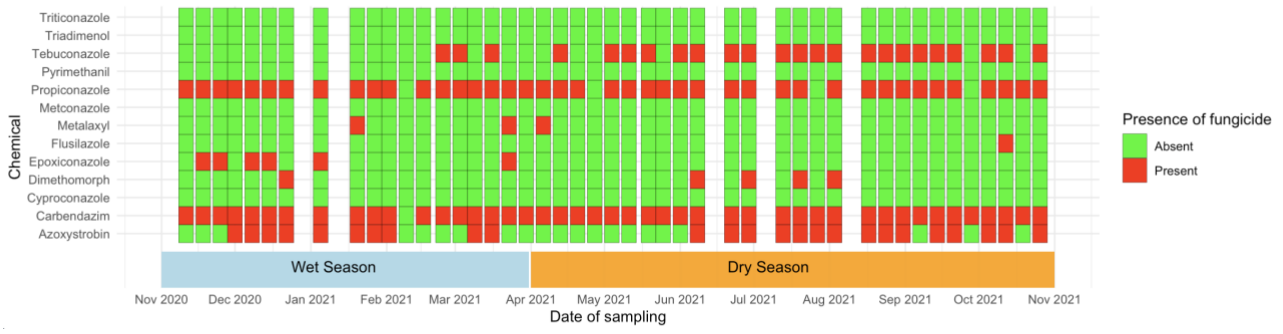

**Figure S2g. PRESENCE AND ABSENCE OF MEDICATIONS OVER A 1-YEAR PERIOD AT SITE 1, STRATIFIED BY MEDICINE CATAGORY**

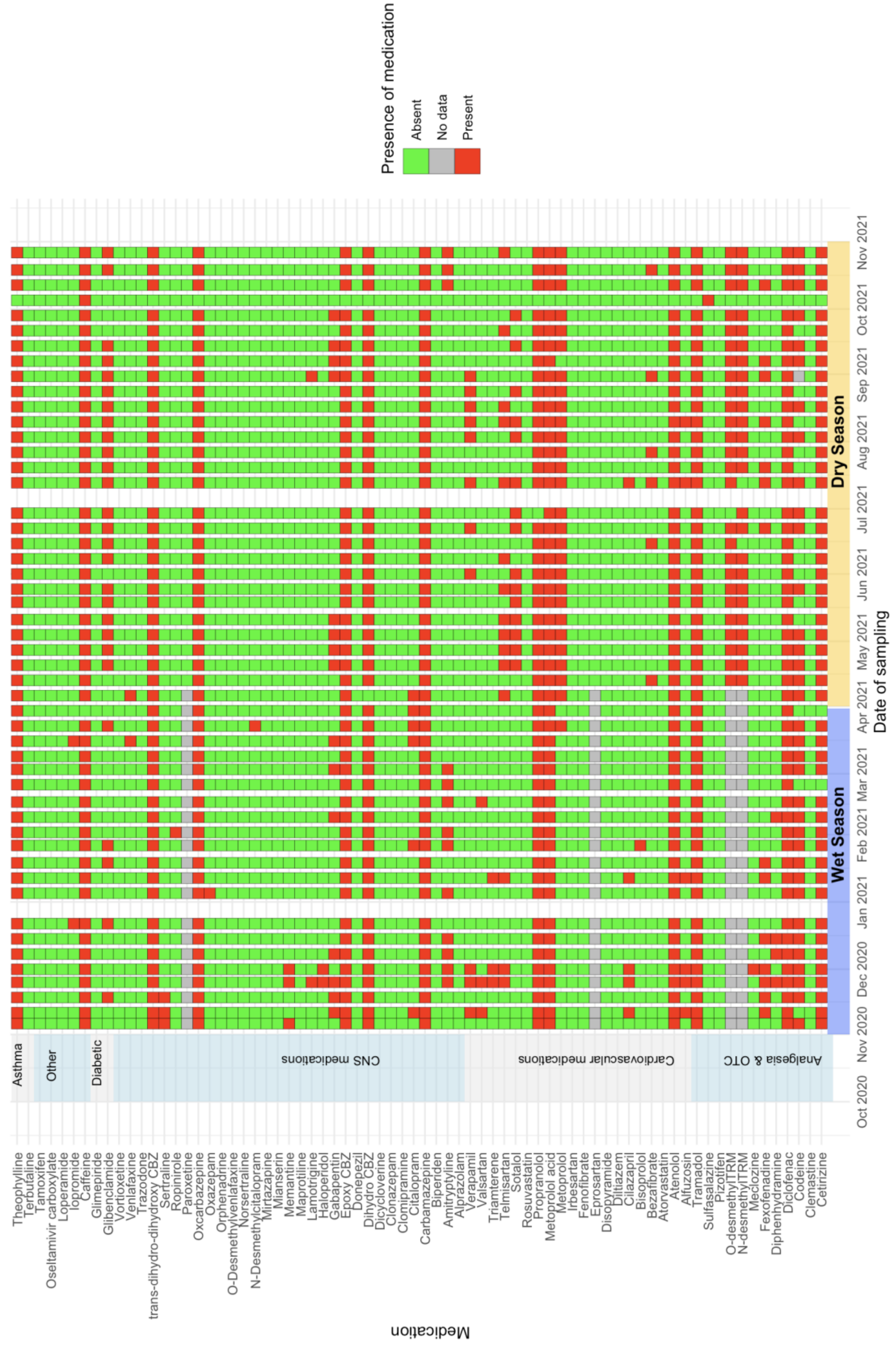

**Figure S2h. PRESENCE AND ABSENCE OF MEDICATIONS OVER A 1-YEAR PERIOD AT SITE 2, STRATIFIED BY MEDICINE CATAGORY**

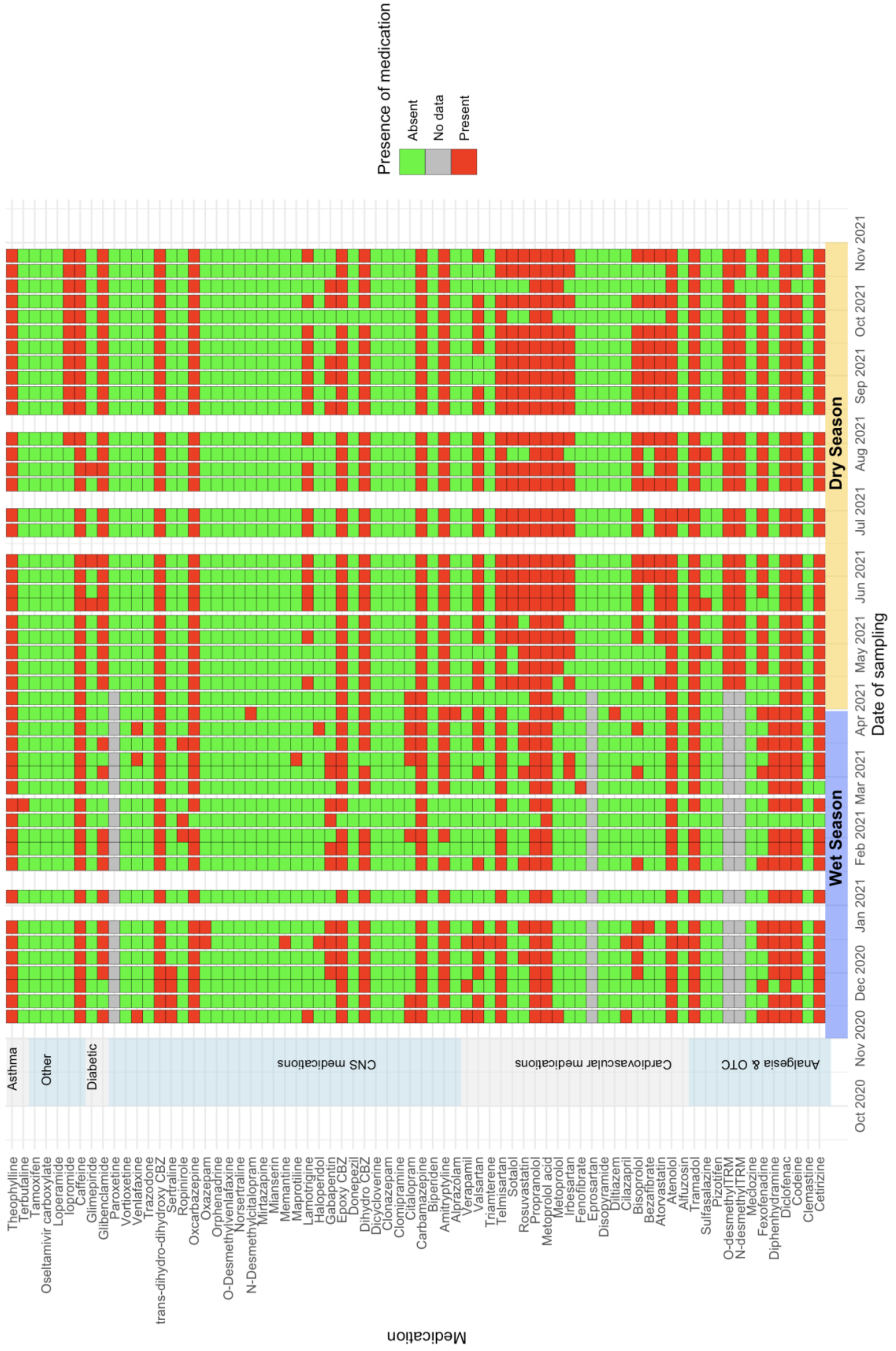

**Figure S2i. PRESENCE AND ABSENCE OF ANTIMICROBIALS OVER A 1-YEAR PERIOD AT SITE 1, STRATIFIED BY ANTIBIOTIC CLASS.**

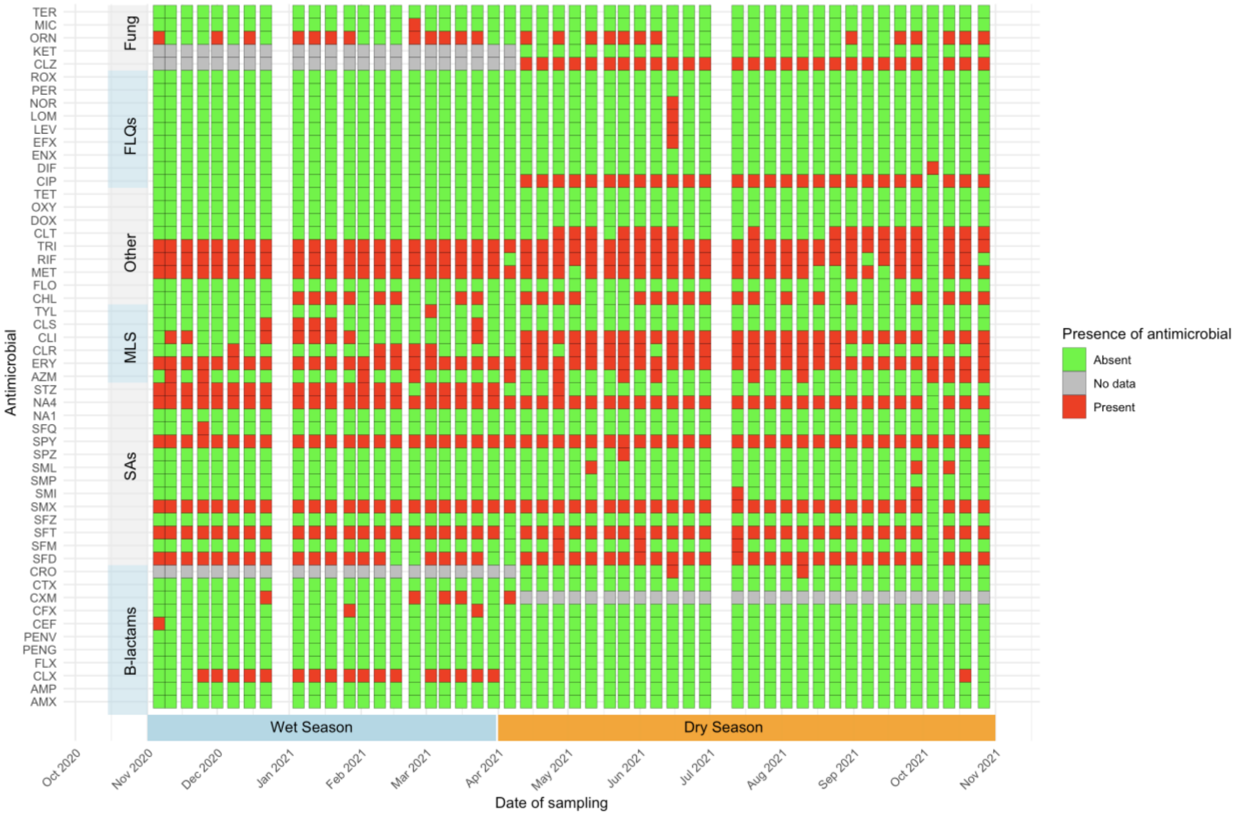


**Figure S2j. PRESENCE AND ABSENCE OF ANTIMICROBIALS OVER A 1-YEAR PERIOD AT SITE 2, STRATIFIED BY ANTIBIOTIC CLASS.**

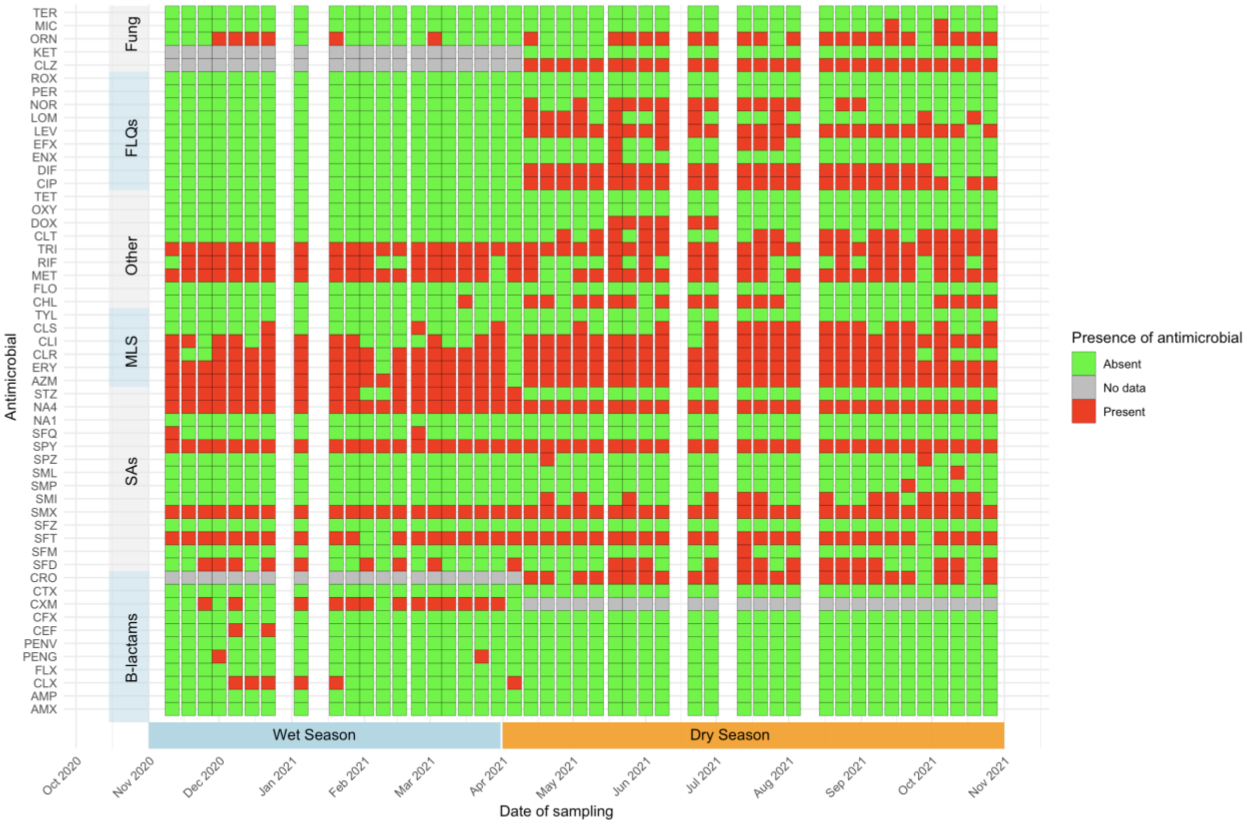

**Figure S3a. SPATIOTEMPORAL VARIATIONS IN THE CHEMICAL COMPOUNDS FOUND AT SITE 1 OVER A 1-YEAR PERIOD.** Presented as the percentage (%) of the total sample amount normalised to sampling time ($ng/POCIS^{-1}/day^{-1}$) representing each chemical class where positive detection of chemicals had been identified.

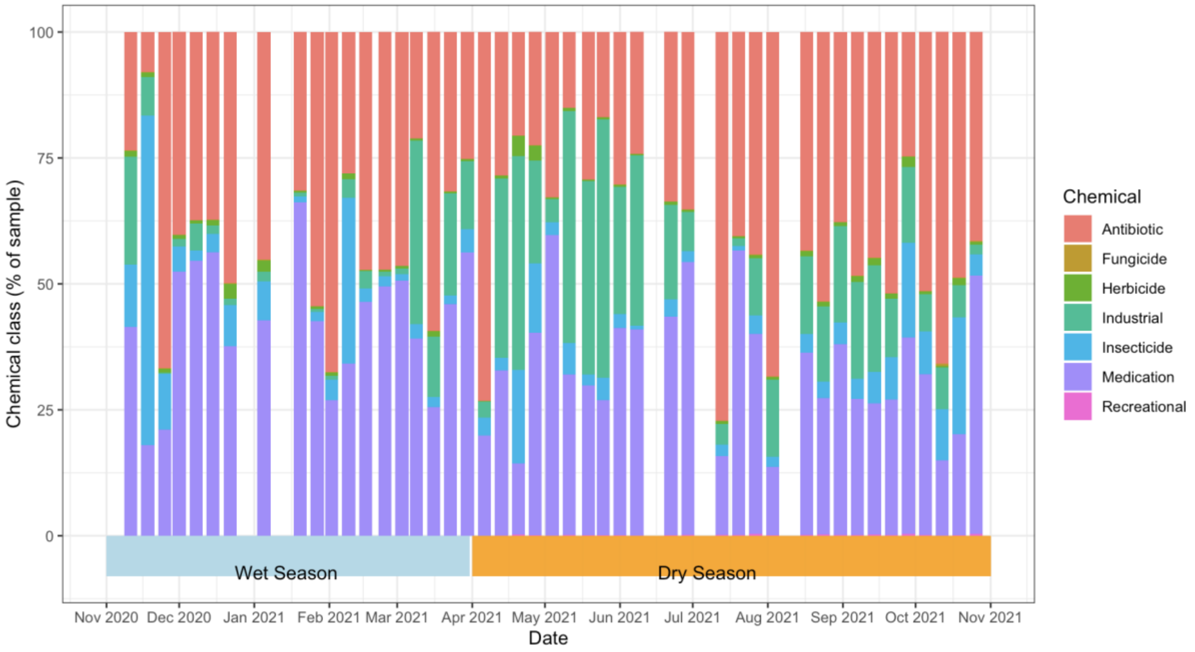


**Figure S3b. SPATIOTEMPORAL VARIATIONS IN THE CHEMICAL COMPOUNDS FOUND AT SITE 2 OVER A 1-YEAR PERIOD.** Presented as the percentage (%) of the total sample amount normalised to sampling time ($ng/POCIS^{-1}/day^{-1}$) representing each chemical class where positive detection of chemicals had been identified.

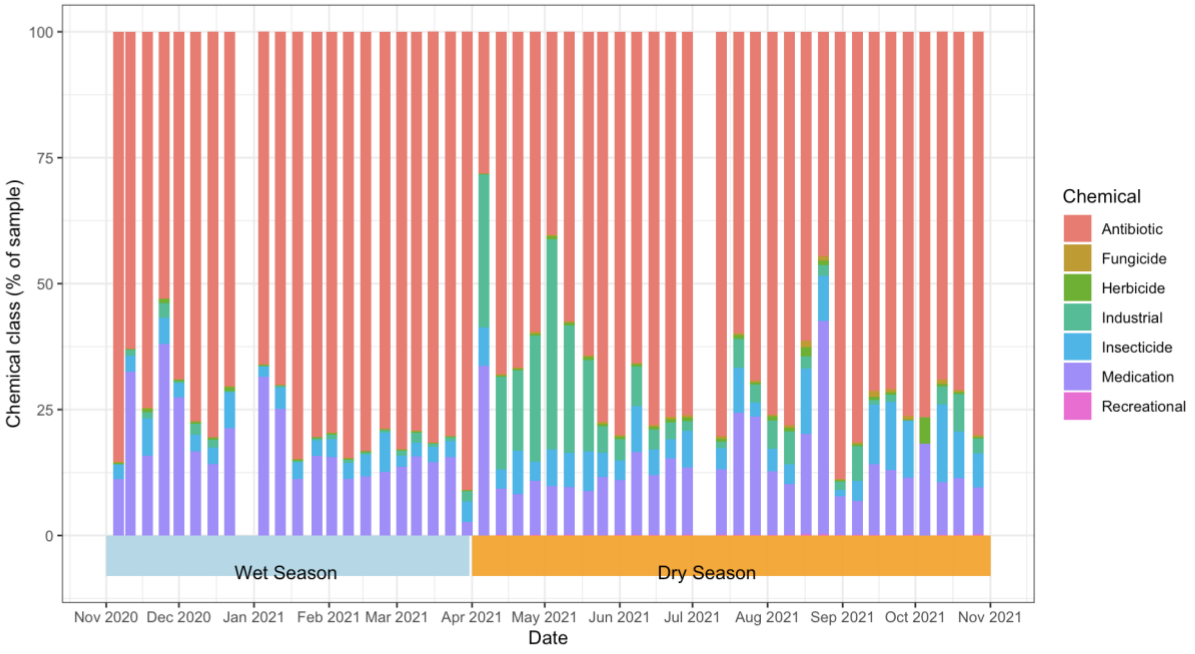

**Figure S4a. SPATIOTEMPORAL VARIATIONS IN MEDICAL COMPOSITIONS FOUND AT SITE 1 OVER A 1-YEAR PERIOD.** Presented as the percentage (%) of the total medication concentration normalised to sampling time (ng/POCIS$^{-1}$/day$^{-1}$) representing each medication.

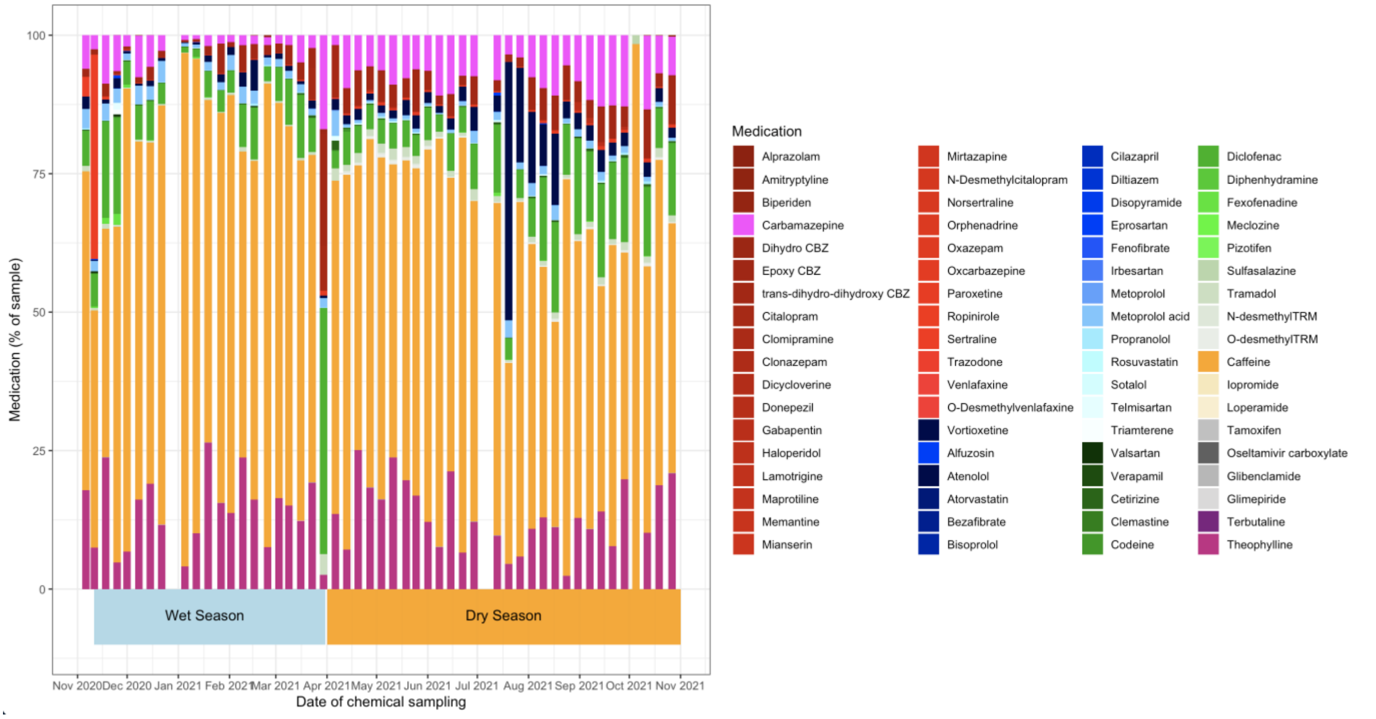


**Figure S4b. SPATIOTEMPORAL VARIATIONS IN MEDICAL COMPOSITIONS FOUND AT SITE 2 OVER A 1-YEAR PERIOD.** Presented as the percentage (%) of the total medication concentration normalised to sampling time (ng/POCIS$^{-1}$/day$^{-1}$), representing each medication.

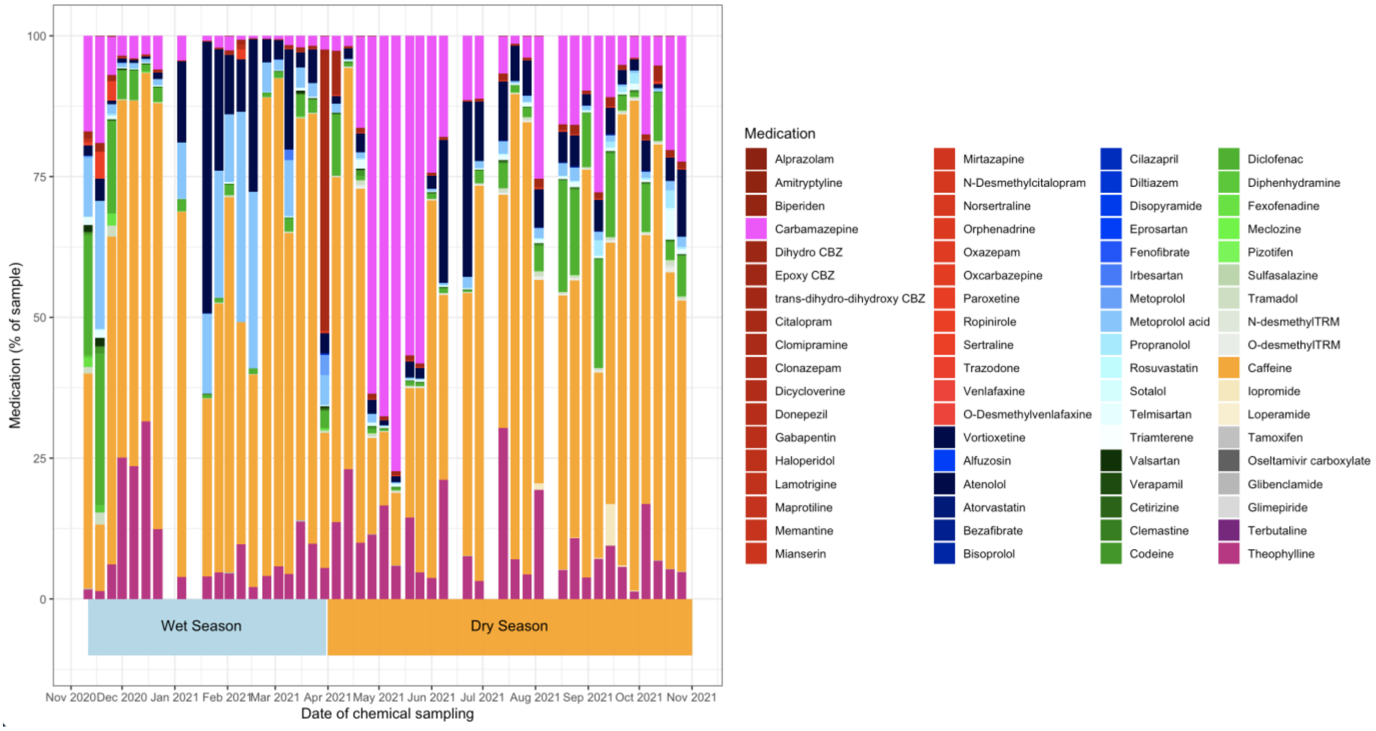

**Figure S5a. SPATIOTEMPORAL VARIATIONS IN ANTIBIOTIC COMPOSITIONS AT SITE 1 OVER A 1-YEAR PERIOD.** Presented as the percentage (%) of the total antibiotic concentration normalised to sampling time (ng/POCIS$^{-1}$/day$^{-1}$) coloured by antibiotic class.

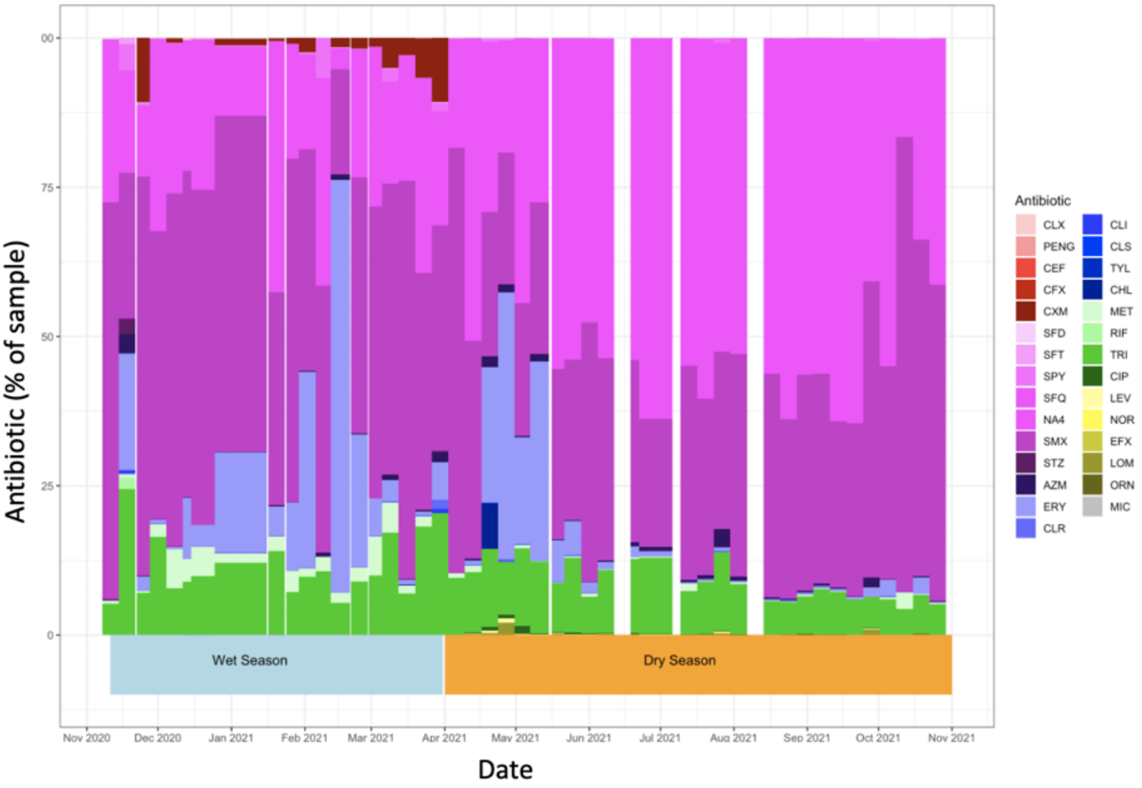


**Figure S5b. SPATIOTEMPORAL VARIATIONS IN ANTIBIOTIC COMPOSITIONS AT SITE 2 OVER A 1-YEAR PERIOD.** Presented as the percentage (%) of the total antibiotic concentration normalised to sampling time (ng/POCIS$^{-1}$/day$^{-1}$) coloured by antibiotic class.

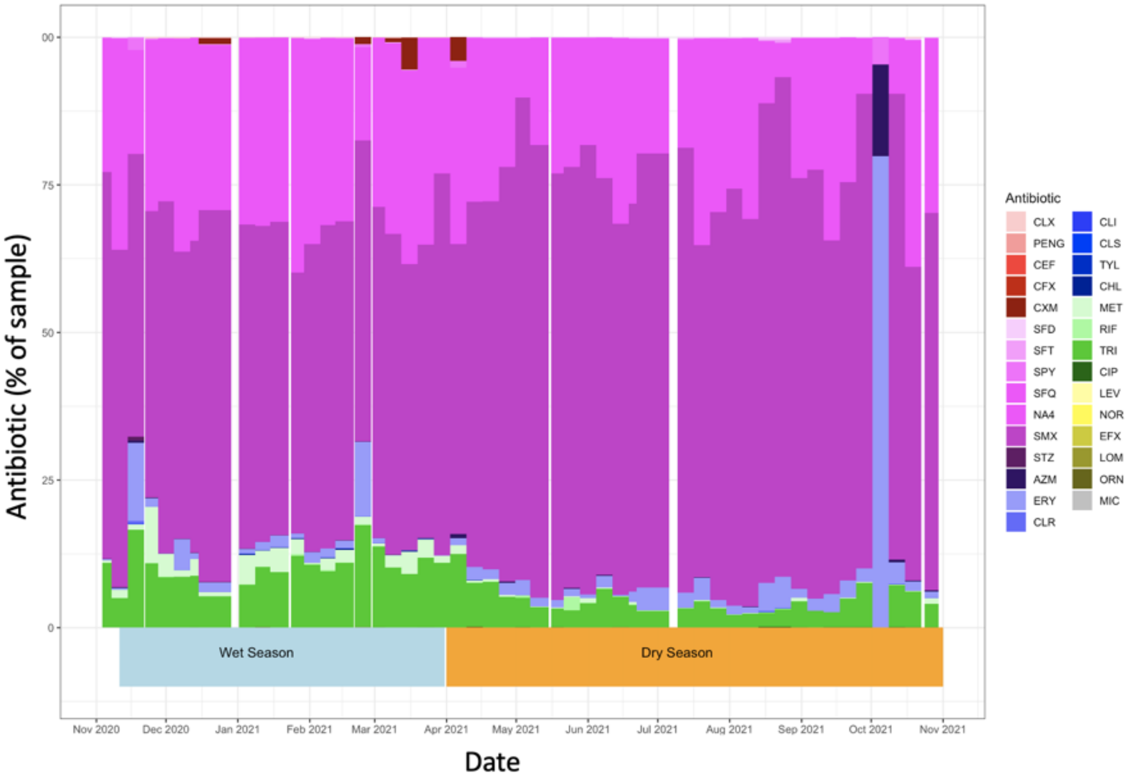

**Figure S6a. VIOLIN PLOT OF INSECTICIDE CONCENTRATIONS (ng/POCIS$^{-1}$/day$^{-1}$) OBTAINED FROM URBAN SITES**.

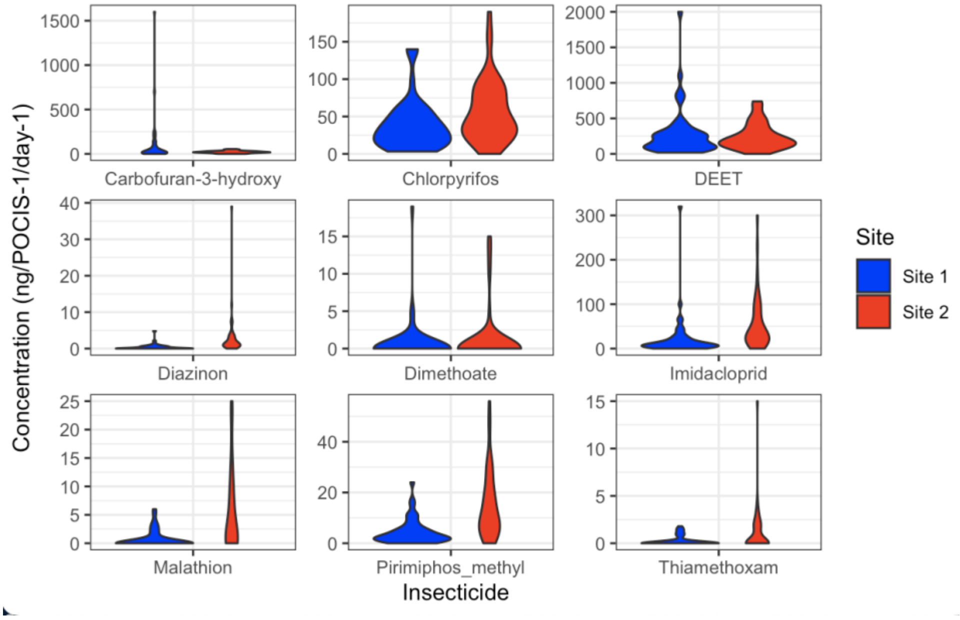


**Figure S6b. VIOLIN PLOT OF HERBICIDE CONCENTRATIONS (ng/POCIS$^{-1}$/day$^{-1}$) OBTAINED FROM URBAN SITES**.

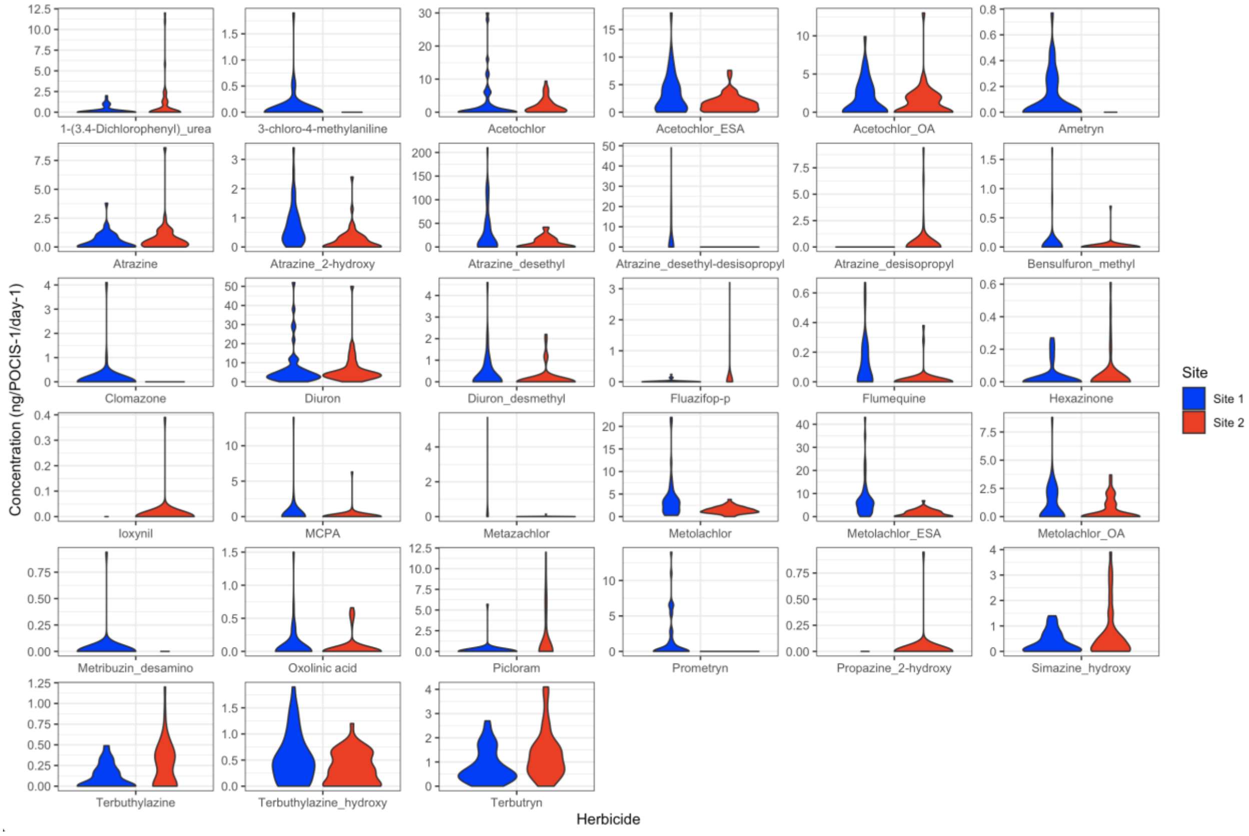

**Figure S6c. VIOLIN PLOT OF FUNGICIDE CONCENTRATIONS (ng/POCIS$^{-1}$/day$^{-1}$) OBTAINED FROM URBAN SITES**.

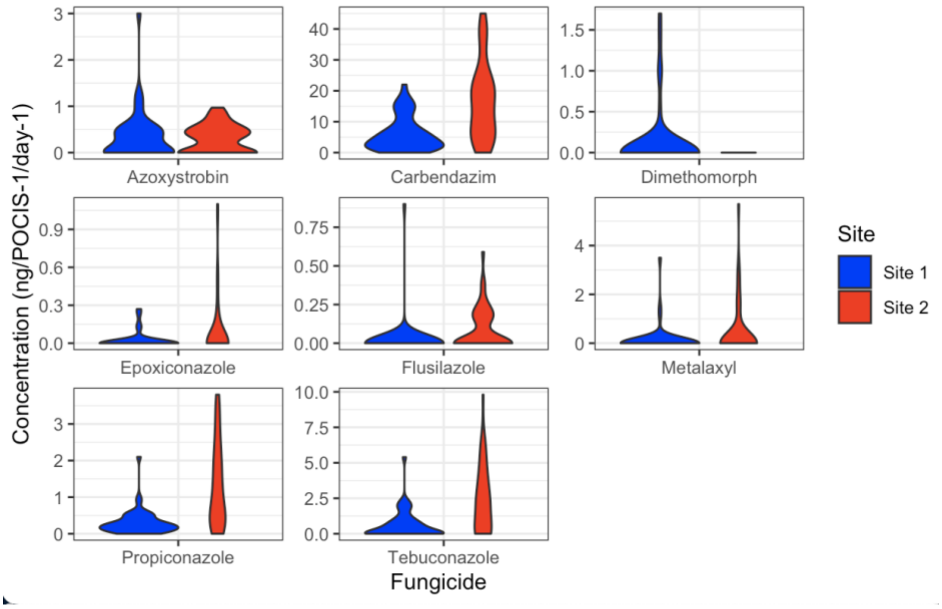


**Figure S6d. VIOLIN PLOT OF INDUSTRIAL CHEMICAL CONCENTRATIONS (ng/POCIS$^{-1}$/day$^{-1}$) OBTAINED FROM URBAN SITES**.

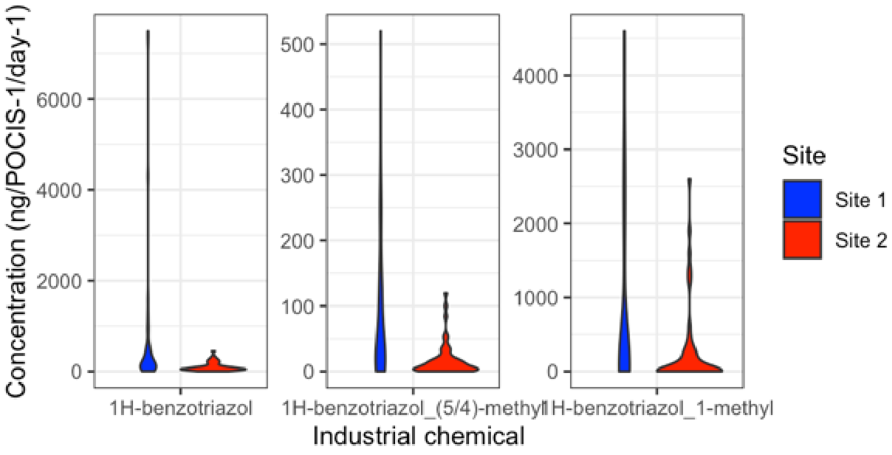

**Figure S6e. VIOLIN PLOT OF RECREATIONAL DRUGS AND ANALGESIA CONCENTRATIONS (ng/POCIS$^{-1}$/day$^{-1}$) OBTAINED FROM URBAN SITES**.

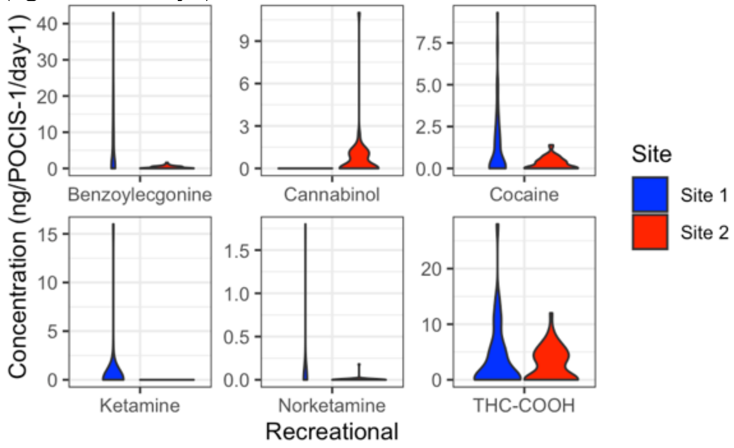


**Figure S6f. VIOLIN PLOT OF HUMAN-USE PHARMACEUTICAL CONCENTRATIONS (ng/POCIS$^{-1}$/day$^{-1}$) OBTAINED FROM URBAN SITES**.

CBZ – carbamazepine, TRM - tramadol

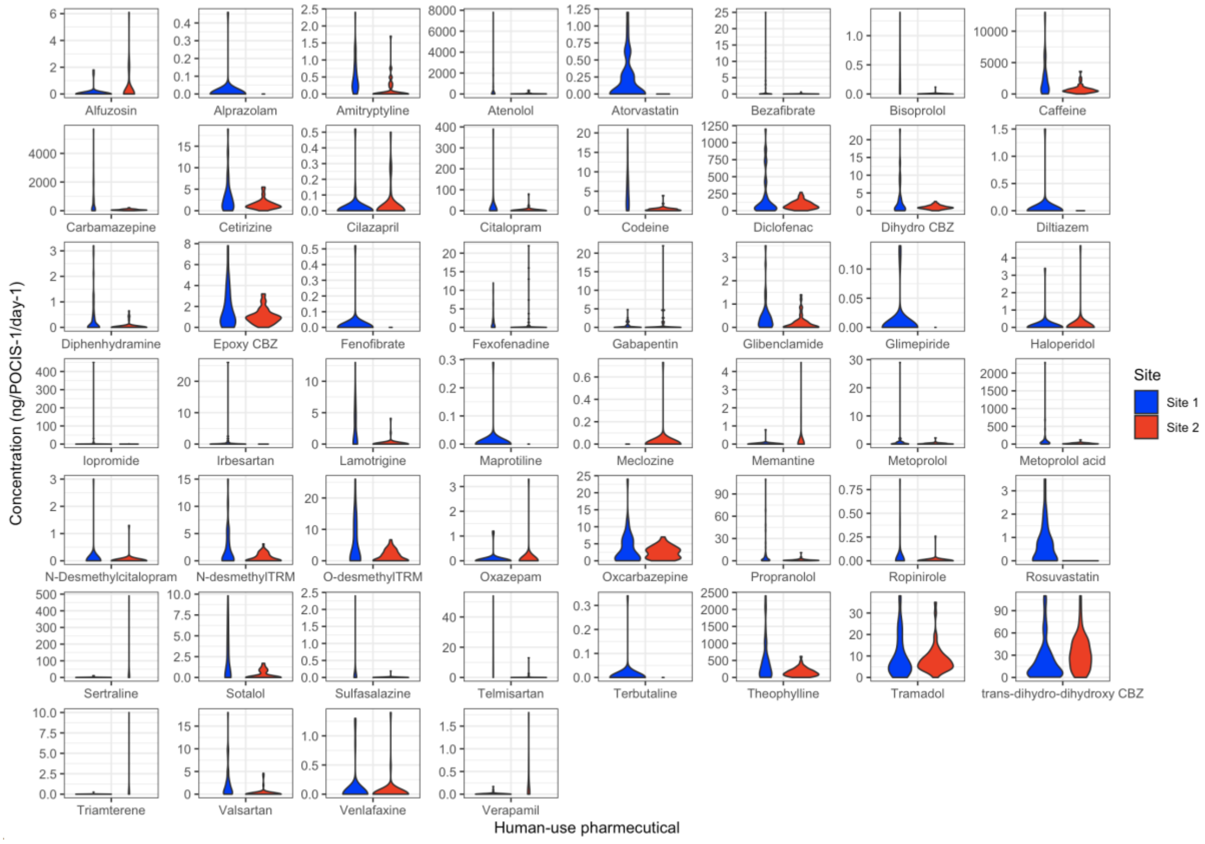

**Figure S6g. VIOLIN PLOT OF ANTIMICROBIAL CONCENTRATIONS (ng/POCIS$^{-1}$/day$^{-1}$) OBTAINED FROM URBAN SITES**.

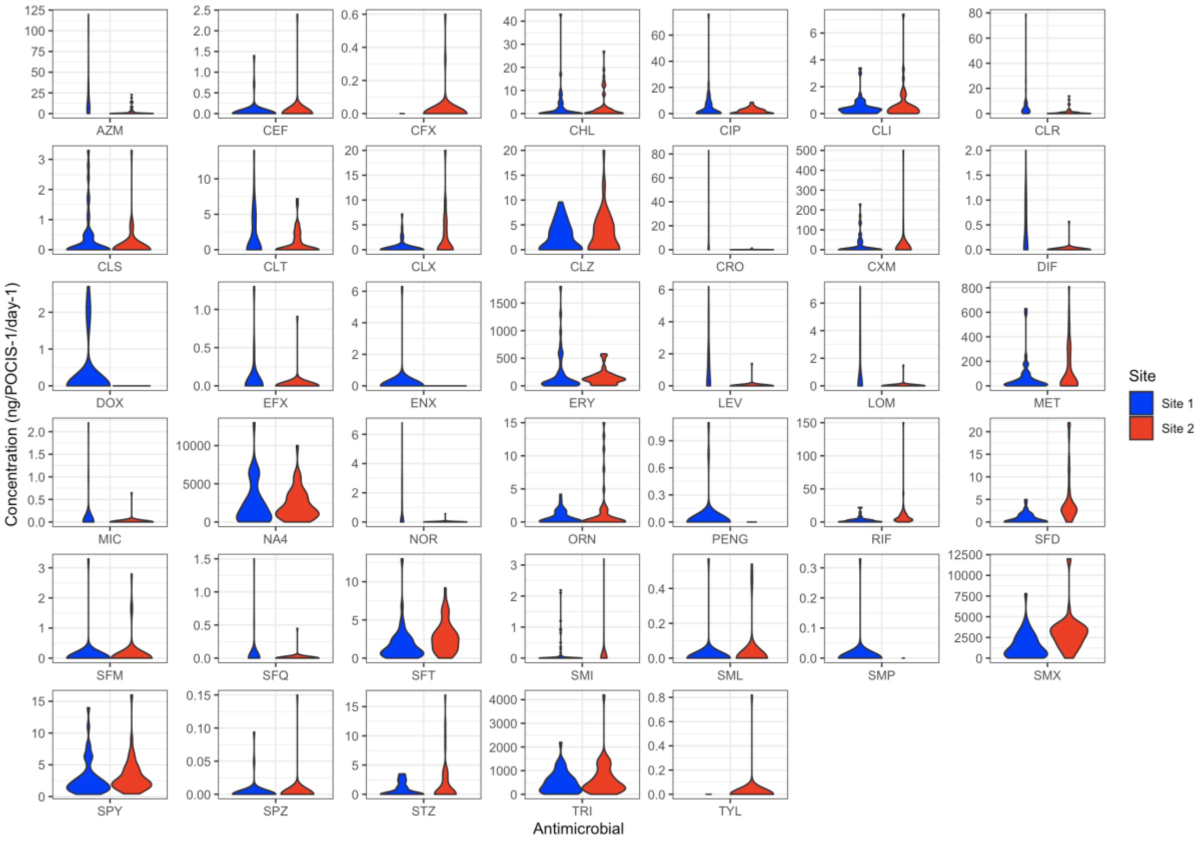


**Figure S7a. CUMULATIVE TOTAL OF MEDICATIONS IDENTIFIED FROM SITE 1, NORMALISED TO SAMPLING TIME (ng/POCIS$^{-1}$/day$^{-1}$), STRATIFIED BY SITE AND COLOURED BY MEDICATION CLASS.**

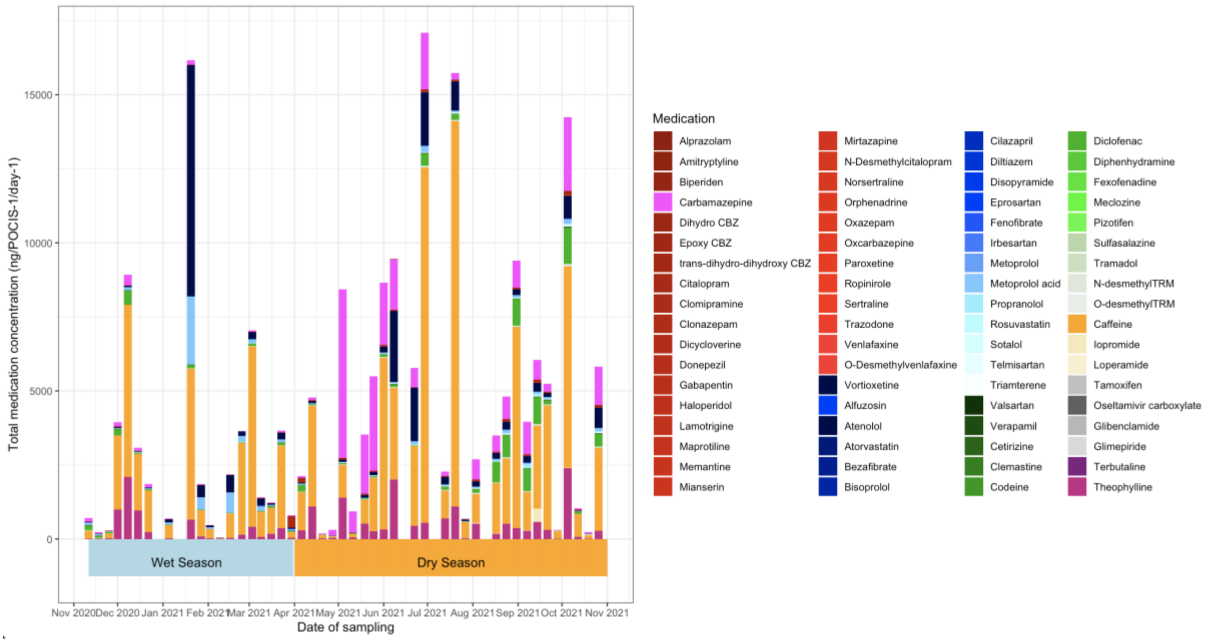

**Figure 7b. CUMULATIVE TOTAL OF MEDICATIONS IDENTIFIED FROM SITE 2, NORMALISED TO SAMPLING TIME (ng/POCIS$^{-1}$/day$^{-1}$), STRATIFIED BY SITE AND COLOURED BY MEDICATION CLASS.**

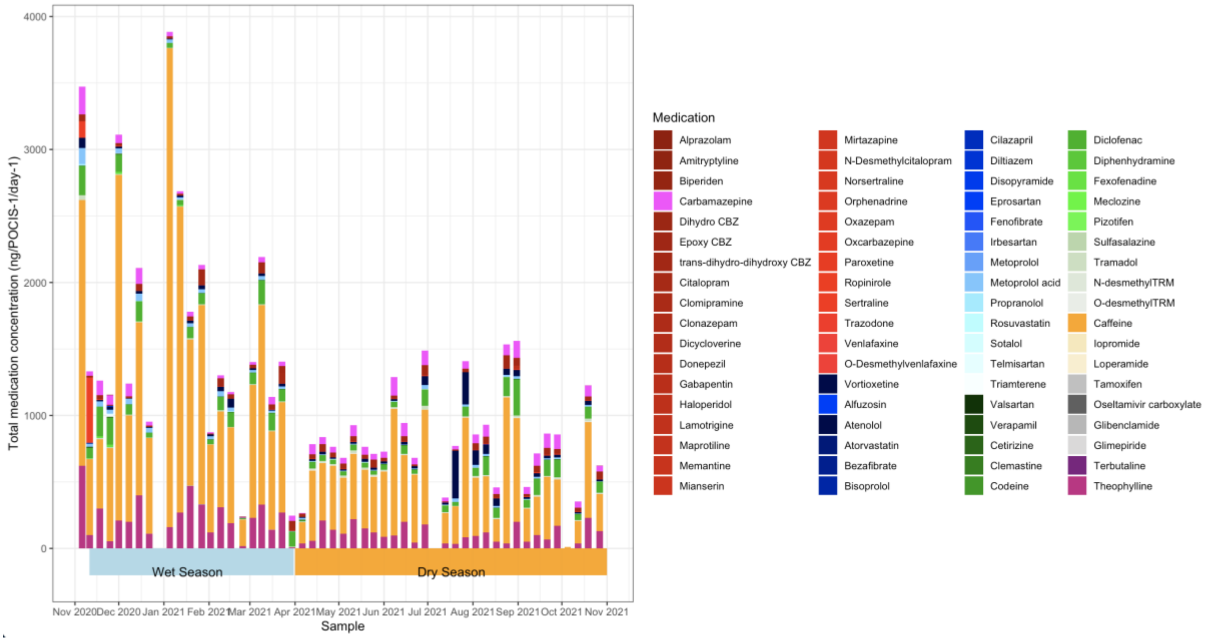


**Figure S8a. TEMPORAL RELATIONSHIPS IN THE RECOVERY AND CONCENTRATION OF MEDICATIONS IN RIVER WATER FROM SITE 1, STRATIFIED INTO SAFE AND UNSAFE PNEC/ CEC LEVELS.**

Monthly trends in the presence and absence (white) of antibiotics are plotted over a 1-year period, spanning across the wet (blue bar) and dry (yellow bar) season. Medications have been stratified into safe (green, <PNEC/CEC) and unsafe (red, >PNEC/CEC) levels based on the concentrations identified. Values inside the cells describe the ratio of analyte:PNEC/CEC illustrating the levels of risk. A value of 0 denotes where an antibiotic was identified above the LOQ but below 0.01% of the agreed PNEC/CEC target.

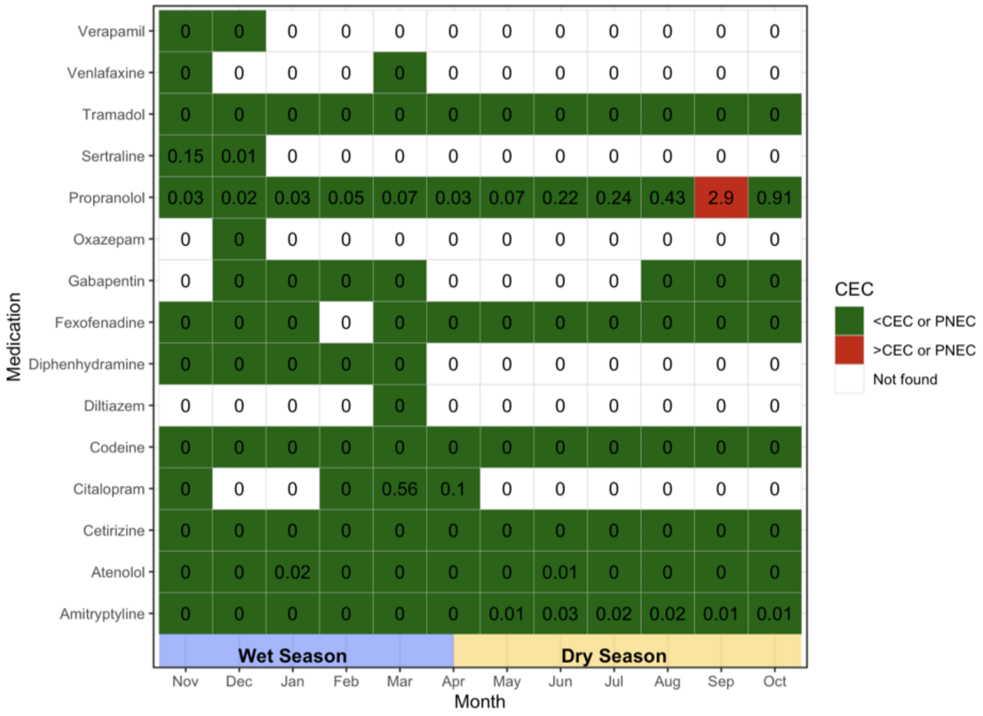

**Figure S8b. TEMPORAL RELATIONSHIPS IN THE RECOVERY AND CONCENTRATION OF MEDICATIONS IN RIVER WATER FROM SITE 2, STRATIFIED INTO SAFE AND UNSAFE PNEC/ CEC LEVELS.**
Monthly trends in the presence and absence (white) of antibiotics are plotted over a 1-year period, spanning across the wet (blue bar) and dry (yellow bar) season. Medications have been stratified into safe (green, <PNEC/CEC) and unsafe (red, >PNEC/CEC) levels based on the concentrations identified. Values inside the cells describe the ratio of analyte:PNEC/CEC illustrating the levels of risk. A value of 0 denotes where an antibiotic was identified above the LOQ but below 0.01% of the agreed PNEC/CEC target.

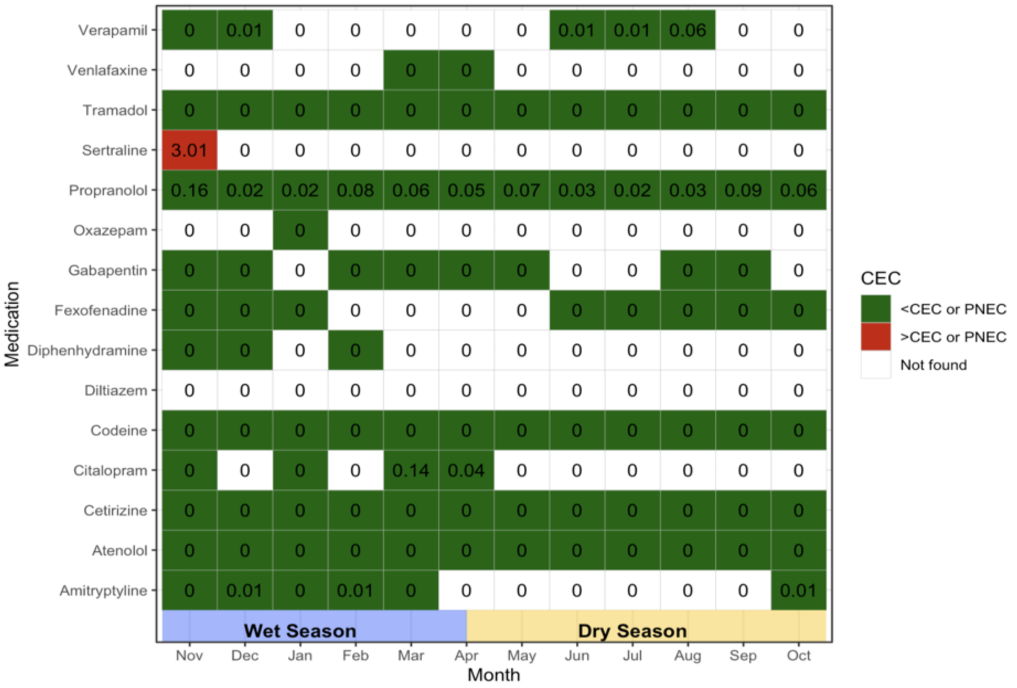


**Figure S9. CUMULATIVE TOTAL OF CHEMICAL COMPOUNDS IDENTIFIED, STRATIFIED BY SITE AND COLOURED BY CHEMICAL CLASS.**

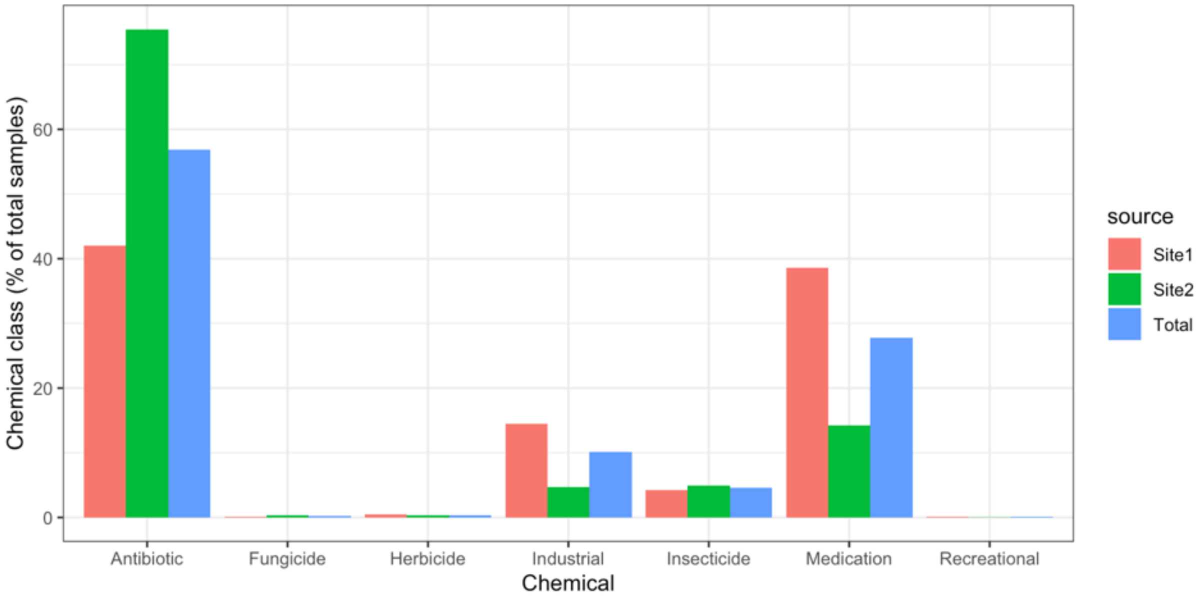

**Figure S10a. CUMULATIVE TOTAL OF ANTIBIOTICS NORMALISED TO SAMPLING TIME (ng/POCIS$^{-1}$/day$^{-1}$) IDENTIFIED FROM SITE 1, STRATIFIED BY SITE AND COLOURED BY ANTIBIOTIC CLASS.** (Wet season = blue background, Dry season = white background).

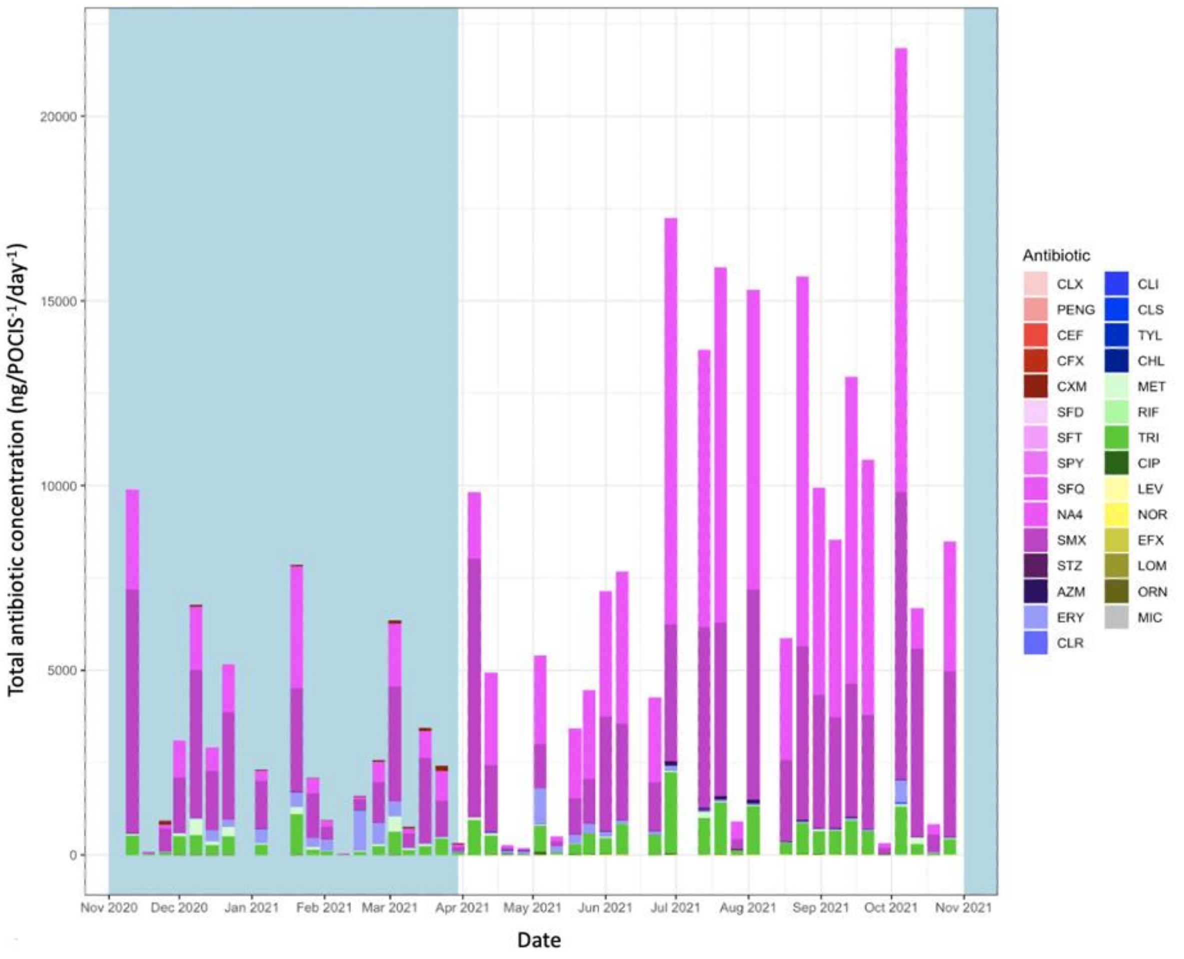

**Figure S10b. CUMULATIVE TOTAL OF ANTIBIOTICS NORMALISED TO SAMPLING TIME ($ng/POCIS^{-1}/day^{-1}$) IDENTIFIED FROM SITE 2, STRATIFIED BY SITE AND COLOURED BY ANTIBIOTIC CLASS.** (Wet season = blue background, Dry season = white background).

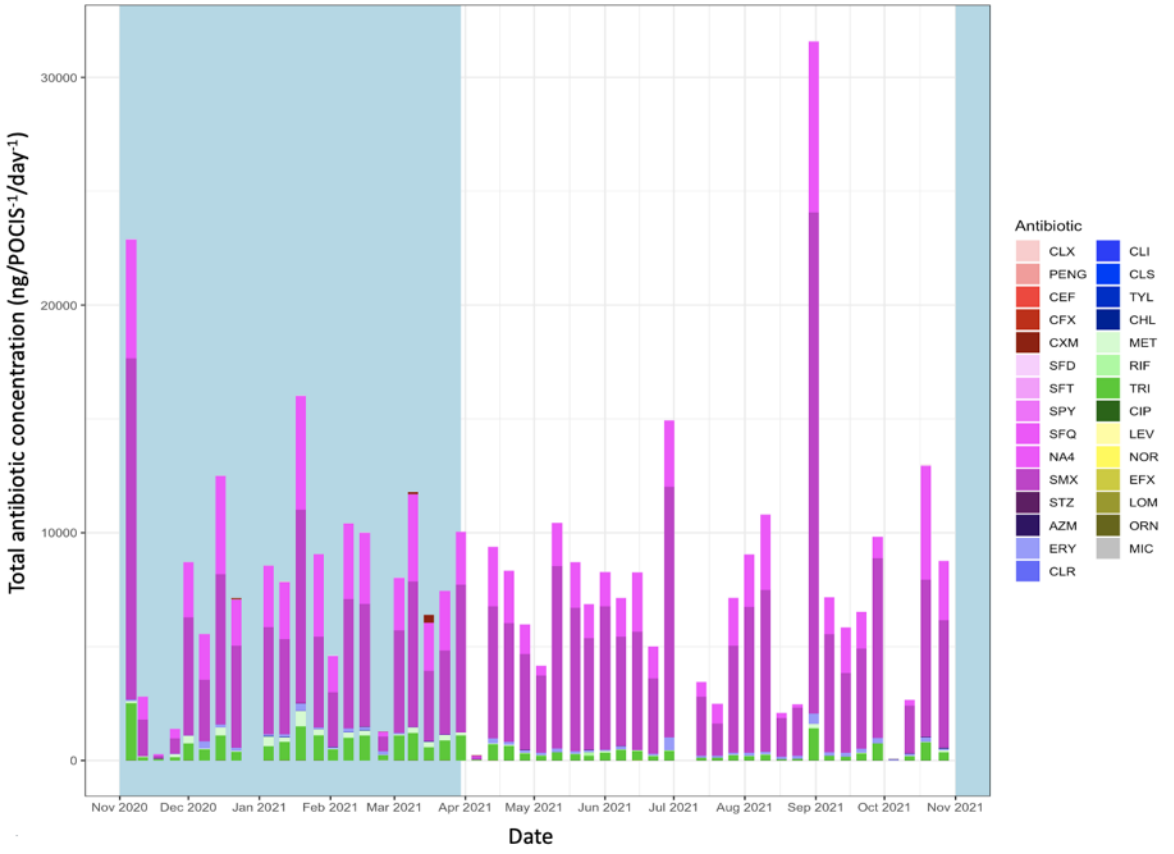


**Figure S11. PEARSONS MATRIX OF ANTIBIOTICS IN RIVER WATER FROM URBAN STUDY SITES.** Correlation coefficients are illustrated on a colour spectrum, with those in red and orange showing the highest degree of relationship.

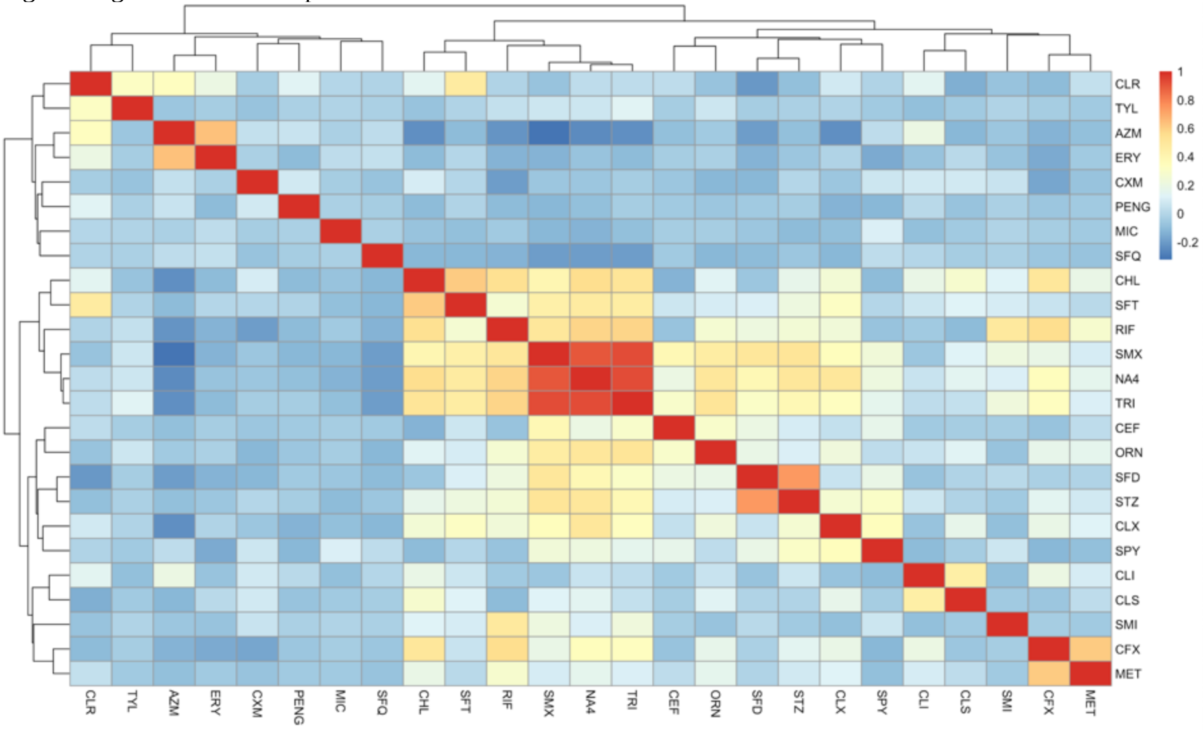

**Figure S12a. TEMPORAL RELATIONSHIPS IN THE RECOVERY AND CONCENTRATIONS OF CUMULATIVE MACROLIDE RISK IN RIVER WATER, STRATIFIED BY SAFE AND UNSAFE PNEC LEVELS FOR EACH SITE.**

Monthly trends in the presence and absence (white) of macrolides are plotted over a 1-year period, spanning across the wet (blue bar) and dry (yellow bar) season. These have been stratified into safe (green, <PNEC) and unsafe (red, >PNEC) levels based on the concentrations identified. Values inside the cells describe the ratio of analyte:PNEC illustrating the levels of risk. A value of 0 denotes where an antibiotic was identified above the LOQ but below 0.01% of the agreed PNEC target. To quantify a cumulative macrolide risk (ng/POCIS$^{-1}$/day$^{-1}$l) we have adjusted each macrolide antibiotic concentration (ng/POCIS$^{-1}$/day$^{-1}$) returned from a sampler compared to a relative PNEC concentrations and added these together, as per the below equation. This will then be compared to the PNEC of azithromycin (20ng/L) to determine if the cumulative risk has exceeded the PNEC level.

$$\begin{aligned}&Modelled\ cummulative\ Macrolide\ risk\\ &= azithromycin\ (ng\ per\ litre) + clarithromycin\ \left(\frac{ng\ per\ litre}{4}\right) + clindamycin\ \left(\frac{ng\ per\ litre}{5}\right)\\ &+ erythromycin\ \left(\frac{ng\ pre\ litre}{25}\right)\end{aligned}$$

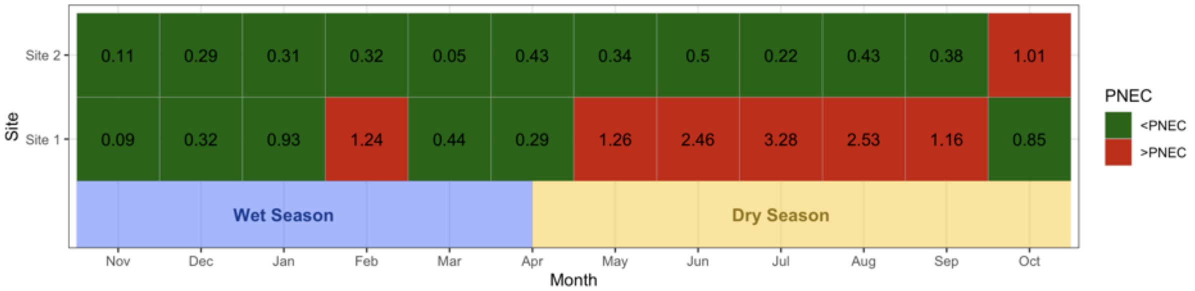


**Figure S12b. TEMPORAL RELATIONSHIPS IN THE RECOVERY AND CONCENTRATIONS OF CUMULATIVE FLUOROQUINOLONE RISK IN RIVER WATER, STRATIFIED BY SAFE AND UNSAFE PNEC LEVELS FOR EACH SITE.**

Monthly trends in the presence and absence (white) of fluoroquinolones are plotted over a 1-year period, spanning across the wet (blue bar) and dry (yellow bar) season. These have been stratified into safe (green, <PNEC) and unsafe (red, >PNEC) levels based on the concentrations identified. Values inside the cells describe the ratio of analyte:PNEC illustrating the levels of risk. A value of 0 denotes where an antibiotic was identified above the LOQ but below 0.01% of the agreed PNEC target. To quantify a cumulative fluoroquinolone risk (ng/POCIS$^{-1}$/day$^{-1}$) we adjusted each fluoroquinolone antibiotic concentration (ng/POCIS$^{-1}$/day$^{-1}$) returned from a sampler compared to a relative PNEC concentrations and added these together, as per the below equation. This will then be compared to the PNEC of ciprofloxacin (60ng/L) to determine if the cumulative risk has exceeded the PNEC level.

$$\begin{aligned}&Modelled\ cummulative\ FQ\ risk\\ &= ciprofloxacin\ (ng\ per\ litre) + enrofloxacin\ (ng\ per\ litre) + flumequine\ \left(\frac{ng\ per\ litre}{4.17}\right)\\ &+ levofloxacin\ \left(\frac{ng\ per\ litre}{4.17}\right) + Norfloxacin\ \left(\frac{ng\ pre\ litre}{8.33}\right)\end{aligned}$$

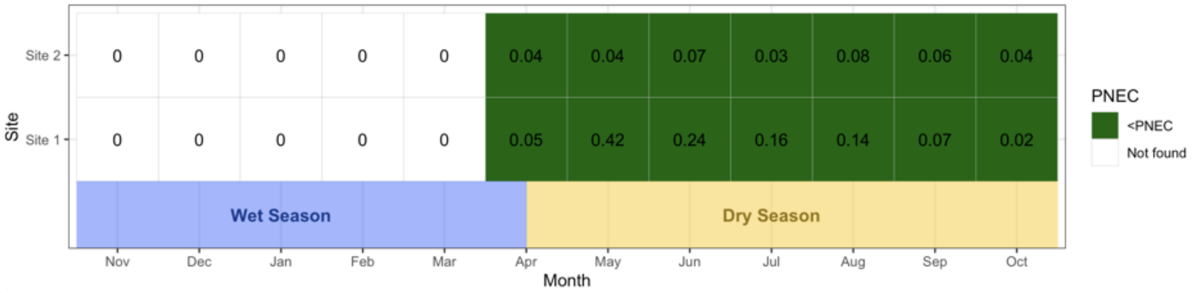

**Figure S13. DETAILED MAPS OF THE RIVERINE NETWORK OF BLANTYRE, INCLUDING (a) BLANTYRE CITY (b) NDIRANDE, AND (c) CHILEKA.**

DRUM study polygons have been demarcated in orange. Sampling sites have been geolocated (site 1: star, site 2: triangle, site 3: square, site 4: circle, site 5: diamond) alongside the key rivers (black = Mudi river, red = Nasolo river, blue = unnamed river).

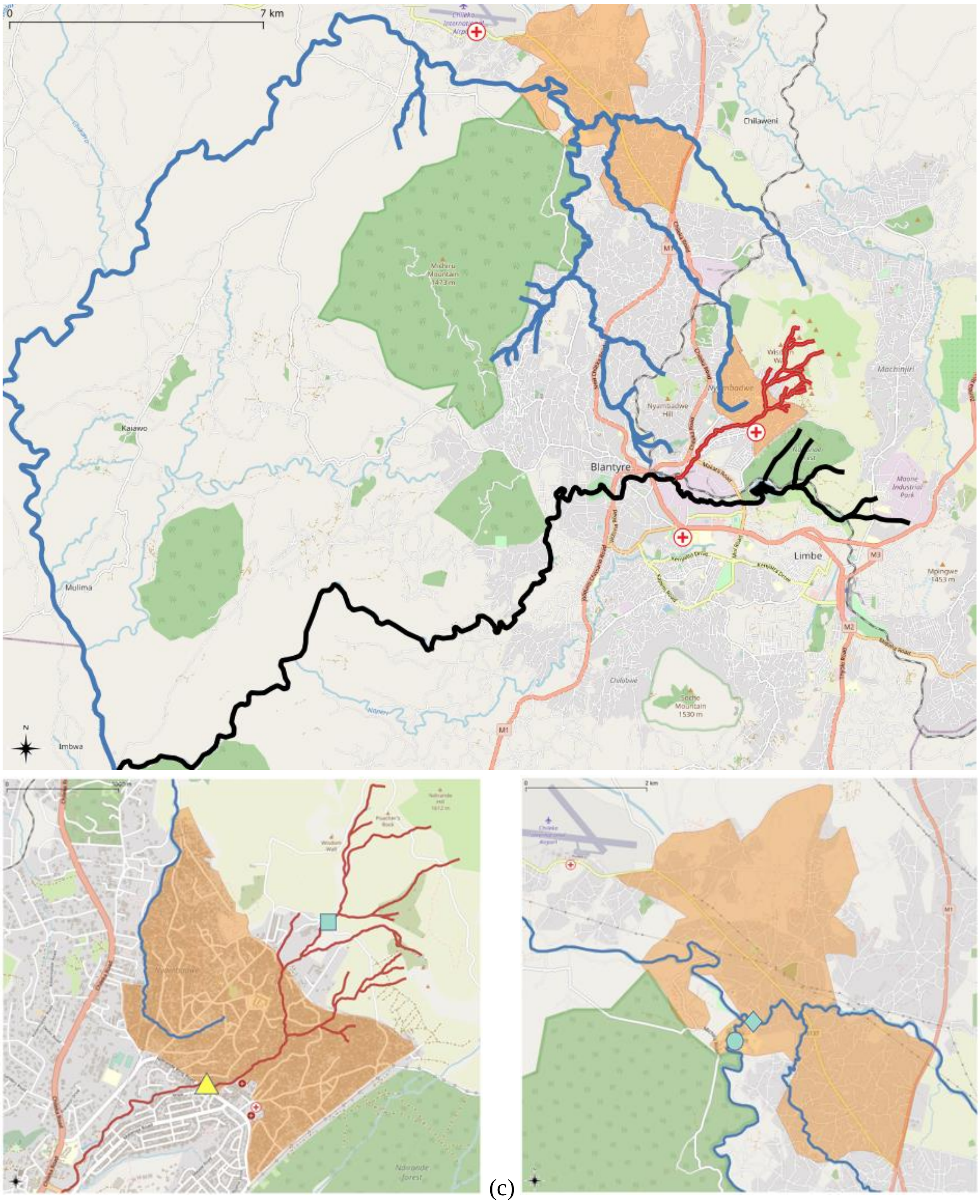


(a)

(b)

(c)

**Figure S14. PHOTOS OF THE SAMPLING LOCATIONS, INCLUDING BOTH STUDY (1&2) AND PILOT (3,4 &5) SITES.** Local approvals and permissions were granted.

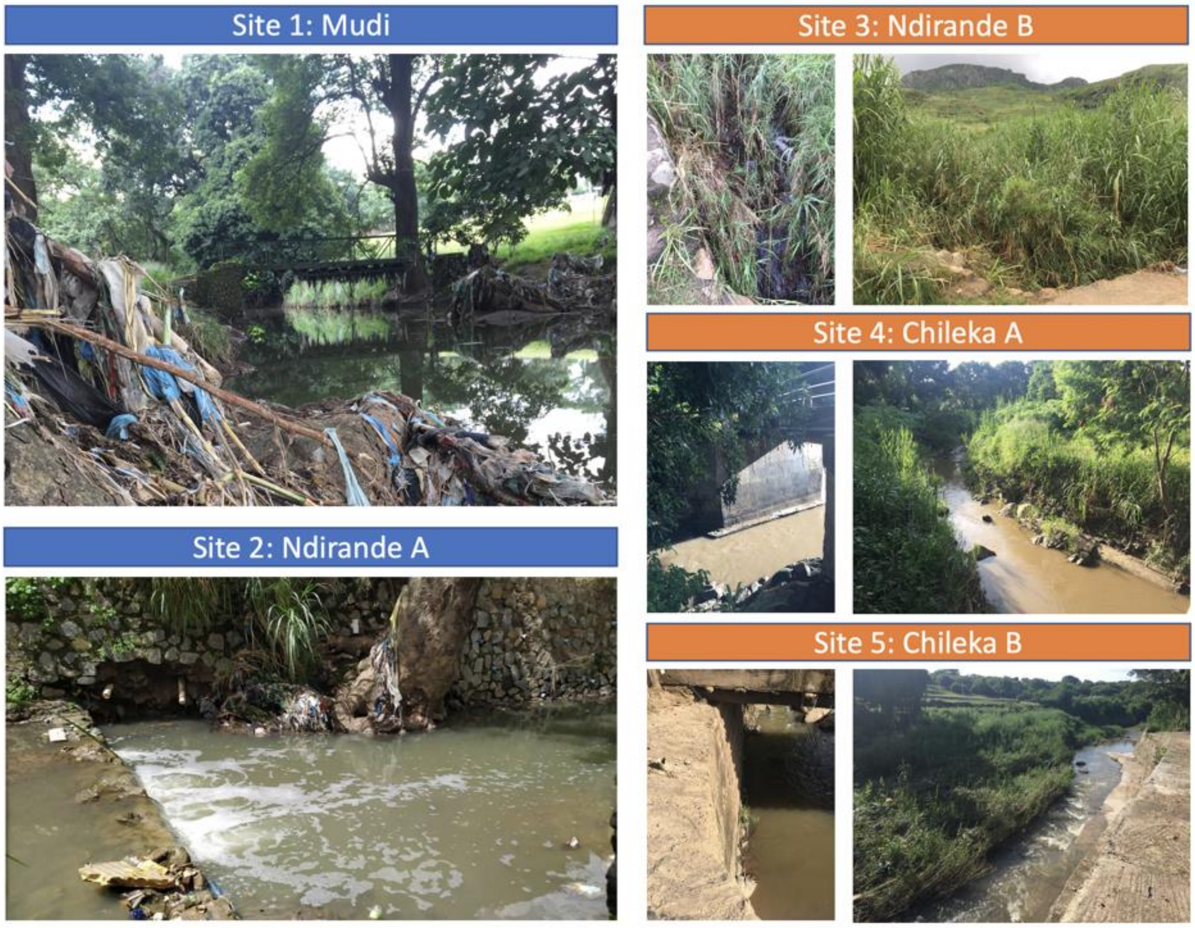

**Figure S15. SEASONAL CHANGES IN THE RIVERS AT SAMPLING SITES**. Photos were taken during the pilot and continuous phase, after approvals and local permissions were granted.

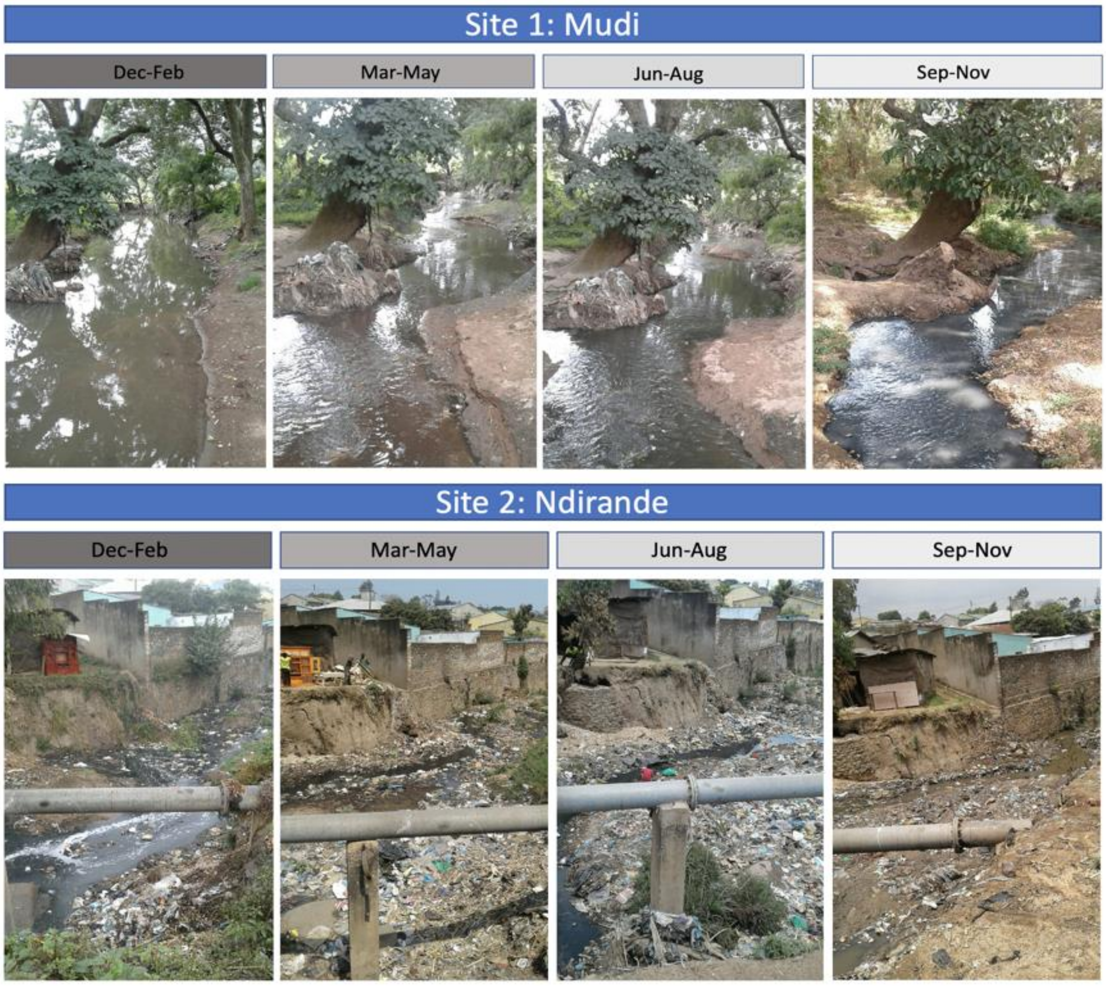


**Figure S16. POCIS SAMPLER**
Porous metal cage (a) sandwiches the PES membrane (b) allowing for environmental exposure while protecting the membrane integrity, which is attached to a metal wire that is secured to the river bank (c).

a) outside cage　　b) internal PES membrane.　　c) completed POCIS with wire

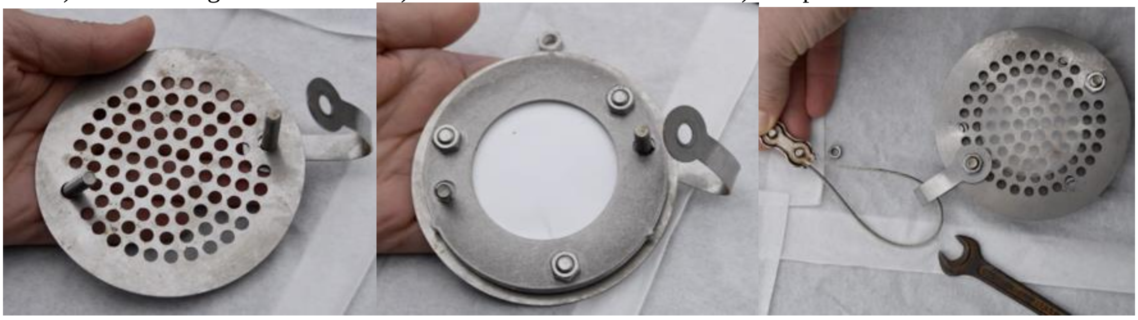

Pictures taken from reference 24 (*Guide for the installation of POCIS passive sampler. Faculty of Fisheries and Protection of Waters, University of South Bohemia, Czech Republic*).
